# Structural epitope profiling identifies antibodies associated with critical COVID-19 and long COVID

**DOI:** 10.1101/2022.07.11.22277368

**Authors:** Patrick K.A. Kearns, Charles Dixon, Mihaly Badonyi, Kim Lee, Rafal Czapiewski, Olivia Fleming, Prajitha Nadukkandy, Lukas Gerasimivicius, Rinal Sahputra, Bethany Potts, Sam Benton, Jacky Guy, Scott Neilson, Helen Wise, Sara Jenks, Kate Templeton, CIRCO, Christina Dold, Teresa Lambe, Andrew Pollard, Alexander J Mentzer, Julian C Knight, COMBAT, Susanna Dunachie, Paul Klenerman, Eleanor Barnes, Alan Carson, Laura McWhirter, Tracy Hussell, Rennos Fragkoudis, Susan Rosser, David Cavanagh, Graeme Cowan, Madhvi Menon, Joseph A. Marsh, Dirk A. Kleinjan, Nick Gilbert

## Abstract

Even within a single protein, antibody binding can have beneficial, neutral, or harmful effects during the response to infection. Resolving a polyclonal antibody repertoire across a pathogen’s proteome to specific epitopes may therefore explain much of the heterogeneity in susceptibility to infectious disease. However, the three-dimensional nature of antibody-epitope interactions makes the discovery of non-obvious targets challenging. We implemented a novel computational method and synthetic biology pipeline for identifying epitopes that are functionally important in the SARS-CoV-2 proteome and identified an IgM-dominant response to an exposed Membrane protein epitope which to our knowledge is the strongest correlate of severe disease identified to date (adjusted OR 72.14, 95% CI: 9.71 – 1300.15), stronger even than the exponential association of severe disease with age. We also identify persistence (> 2 years) of this IgM response in individuals with longCOVID, and a correlation with fatigue and depression symptom burden. The repetitive arrangement of this epitope and the pattern of isotype class switching is consistent with this being a previously unrecognized T independent antigen. These findings point to a coronavirus host-pathogen interaction characteristic of severe virus driven immune pathology. This epitope is a promising vaccine and therapeutic target as it is highly conserved through SARS-CoV-2 variant evolution in humans to date and in related coronaviruses (e.g. SARS-CoV), showing far less evolutionary plasticity than targets on the Spike protein. This provides a promising biomarker for longCOVID and a target to complement Spike-directed vaccination which could broaden humoral protection from severe or persistent disease or novel coronavirus spillovers.

**One-Sentence Summary:** Using a novel protein-structure-based B cell epitope discovery method with a wide range of possible applications, we have identified a simple to measure host-pathogen antibody signature associated with severe COVID-19 and longCOVID and suggest the viral Membrane protein contains an epitope that acts as a T independent antigen during infection triggering extrafollicular B cell activation.

## Introduction

B cells are selected for the affinity of their membrane-bound immunoglobulin receptor (mIg) to bind surfaces on foreign materials, for example, viral proteins. Cells are activated by binding cognate antigen and within a suitable co-stimulatory environment are expanded and differentiate to secrete matching antibodies into blood and mucosa (*1*). Anti-viral antibody binding may be functionally useful, for example, neutralising a microbe; neutral; or harmful, for example, by causing antibody dependent enhancement (ADE) or autoimmunity (*2, 3*)
. Individuals acquire remarkably heterogeneous repertoires of antibodies after encountering a pathogen (*4*).

The heterogeneity in the repertoires and in the functional effects of specific antibodies can therefore determine susceptibility to infection or disease (*5*). However, the affinity of antibodies for three-dimensional structures (epitopes) in conformationally complex large proteins makes deconvoluting the polyclonal B cell response to specific targets a technical challenge (*6, 7*)
. T cells on the other hand inherently recognize digested proteins as peptides presented on MHC molecules, and so can be comprehensively screened in an unbiased way using peptides. Each pathogen protein typically comprises hundreds or thousands of three dimensional (3D) B cell epitopes, such that an assay against a whole conformationally intact protein is an aggregate of the polyclonal response across many targets. Therefore functionally important sites can be missed in the aggregate if there are harmful and beneficial targets within the same protein or the antibodies of importance are a minor component of the whole response (*8*). This is not unusual, for example neutralizing antibodies are typically a minor component of a response to a single protein and bind regions on viral proteins in proximity to sites where non-neutralising antibodies can bind and compete. Profiling antibody binding at the epitope level is therefore important but laborious and does not scale easily for most of the repertoire.

So-called “linear” B cell epitopes –where a continuous peptide fragment (∼10 amino acids) binds antibody– can be screened for using high-throughput methods thanks to their biochemical simplicity. However, these are a minor subset (∼5-10%) of the whole repertoire (*7*), and most are thought to capture functionally irrelevant antibodies that bind only degraded rather than whole proteins exposed on viable pathogens making them more useful as epidemiological tools than in functional antibody discovery (*6, 7*)
. This leaves the possibility that functionally important antibody targets remain undiscovered in the SARS-CoV-2 proteome despite intense study. An alternative to experimental linear epitope discovery methods are in silico tools which predict the position of discontinuous 3D epitopes within the structure of a protein (*9*). However, to date, none of these identify whether a minimally sufficient set of amino acid residues can recapitulate the predicted native epitopes when expressed in isolation.

To address this, we have developed a computationally efficient method for predicting which parts of a protein can form thermodynamically stable peptides that adopt similar 3D conformations when isolated as peptides as within the full-length protein. These peptides are more likely to bind the same antibodies in diverse settings. Our approach aims to greatly enrich peptides that are good candidates to be functionally important and immunodominant: as they are exposed on whole proteins and are likely to persist in the same shape in debris. Experimental efficiency is improved by 1/ not synthesizing peptides that are unstable and 2/ reducing false positives by eliminating stable peptides that will not adopt their native conformation.

This method was then applied to the SARS-CoV-2 proteome to examine all possible peptides between the arbitrary cutoffs of 10-100 amino acids in length. High-ranking peptides were then experimentally validated by screening antibody repertoires from patients in independent clinical cohorts recruited within the UK National Health Service. Severe SARS-CoV-2 infection is characterised by a strikingly stereotyped acute immunological illness in the period after peak viral load (COVID-19), (*10*) and antibodies are already known to be both a key correlate of protection from infection (*11*) and a cause of harm (*5, 12*)
. However, COVID-19 outcome heterogeneity remains incompletely explained and the virus-specific factors that trigger breakdowns in tolerance and resultant immunopathology for SARS-CoV, MERS-CoV, and SARS-CoV-2 but not for other coronaviruses are not known. We hypothesized that there may be discoverable targets within the SARS-CoV-2 proteome that contribute to dysregulated immunity, by molecular mimicry, antibody dependent enhancement or another mechanism. Pursuing this hypothesis, an epitope was identified that is strongly associated with both acute COVID-19 and longCOVID immunopathology and preliminary data supports a novel host:pathogen interaction which is a promising target for therapeutic intervention.

## Results

### Validation of computational method for predicting structurally-stable epitopes

A 1000 amino acid long protein has 86,086 possible sub-peptides between 10 and 100 residues in length. Across the SARS-CoV-2 proteome there are 1,240,901 such peptides. As it is expensive to screen this many peptides experimentally, or computationally using complex energy functions, we took advantage of a property that is simple to compute from protein structure and is directly related to the energy of protein folding: the solvent-accessible surface area (SASA). Previously, we have demonstrated the utility of SASA for prediction of protein stability, flexibility, and assembly pathways, and shown that it is competitive with much more computationally intensive structural modelling strategies (*13–15*), making it feasible to assess a huge number of possible peptides.

Starting with all proteins from the reference SARS-CoV-2 proteome (uniprot.org/proteomes/UP000464024) (Fig. 1A), we selected proteins where structural models were available, and then fragmented the structures into all possible 10-100 amino acid sub-peptides. For these sub-peptides, we computed a metric we term ΔASA_r_ (which we pronounce “DASA”), defined as the difference in SASA between the free peptide and the peptide in the context of the full protein structure normalised by the SASA of the peptide. Peptides with low ΔASA_r_ make fewer contacts outside of the peptide region and are thus more likely to maintain in the free form a conformation like that in the native structure (Fig. 1B). Applying our ΔASA_r_ approach to external experimental datasets from Phage-display-based antibody screening platforms (“VirScan” (*16*) and “ReScan” (*17*)), we find we are able to predict which of a pair of overlapping peptides within these datasets spanning a region was more antigenic despite neither peptide in these pairs being likely to be as stable as a peptide where the length and position are optimized to minimize ΔASA_r_ (Fig. S1).

**Fig. 1.**
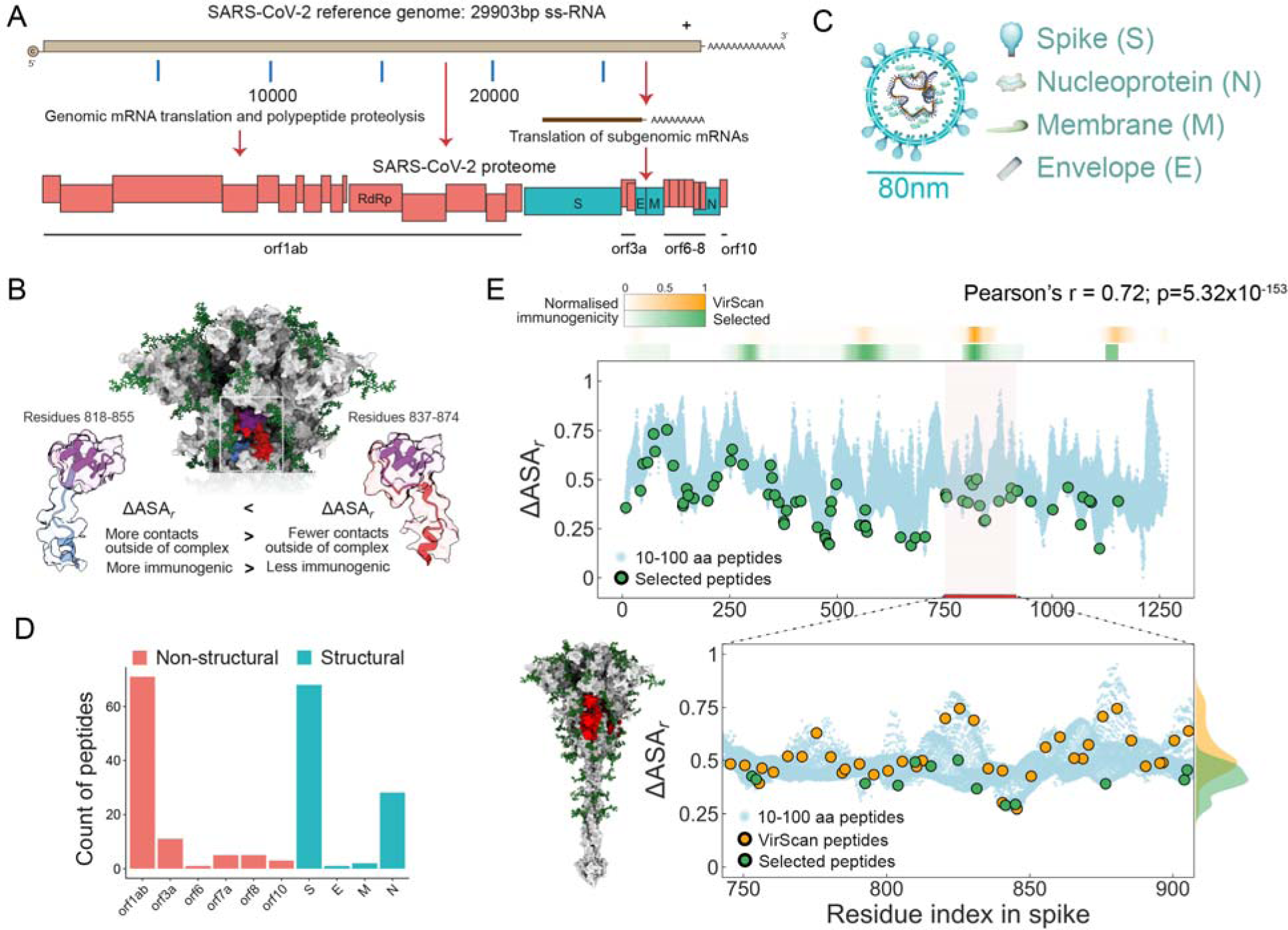
Predicting structurally stable epitopes from the SARS-CoV-2 proteome. (A) SARS-CoV-2 reference genome and proteome. (Red – non-structural proteins; Cyan – structural proteins). (B) Peptides with lower ΔASA_r_ are more likely to adopt similar conformations as free peptides and are hypothesized to be more immunogenic than peptides covering the same region having higher ΔASA_r_. (C) The structural proteins of a SARS-CoV-2 virion. (D) Count of the 196 peptides from SARS-CoV-2 proteome by viral protein. (E) Top: the blue dots show the values of ΔASA_r_ for all possible 10-100 residue long peptides in spike along the linear sequence. Peptides are represented by their midpoints. Green dots mark the selected peptides using our structure-guided approach. Bars on top are co-linear with the residue index and show the immunogenic profile of spike as determined by VirScan phage-display (orange) or by our approach (green). VirScan: Z-score difference, ours: ELISA ratio of positive sera to negative sera smoothened with a sliding window average of +/- 10aa and normalized to a scale of 0-1. Without using the sliding window average the Pearson’s correlation is r = 0.68 (p=8.21×10^-129^). Bottom: the amino acid region 750-900 of spike is shown in the structural context (left) or as a ΔASA_r_ distribution (right). Orange dots are midpoints of VirScan peptides, which generally have higher ΔASA_r_ values, as highlighted by the density diagram on the right (p<1×10^-3^ comparing all Spike proteins for both methods, Wilcoxon two-sided rank sum test).

Although of two antibody-binding peptides, we expect the more stable peptide to be a better immunogen, all else being equal, our stability-prioritizing strategy was not developed to predict immunogenic regions of the proteins, per se. Consequently, to experimentally validate our method, we next prioritized the stable peptides from proteins we judged most likely to be functionally relevant. For example, we prioritized peptides from the virion structural proteins, as these are the most abundant antigens in the extracellular environment. In total, we selected 100 peptides from the structural proteins (Fig. 1C), and a range of peptide sizes and selected 96 peptides from the non-structural proteins (Fig. 1D), but without prioritisation using ΔASA_r_ values as structural models were not available at the outset of this project (April 2020).

We designed expression vectors for high-throughput, robot-assisted cloning and used these to individually express and purify our predicted stable peptides as GST-fusion proteins. Almost all (98%, 192/196) predicted stable peptides were successfully expressed in a bacterial expression system. Isotype-specific reactivities in sera were then quantified by ELISA (Fig. S2).

Pooled sera (3M; Technopath, Tipperary, Ireland) from SARS-CoV-2 recovered and pre-2019 naïve subjects was used to validate the reactivity of the peptides. Figure 1E, demonstrates a general overlap between our method and the phage-display method “VirScan” (Pearson’s r=0.72, p=5.32×10^-153^) for the Spike protein, but our method identified stronger immunogenicity at some of these regions (*16*). The VirScan library uses arbitrarily sized k-mers, with arbitrarily sized overlaps, which are often not structurally optimal, as judged by the ΔASAr (Fig. 1E) (*16*).

Consistent with other studies (*16–18*), most immunodominant epitopes were found within S and N (Fig. 2A & B, Fig. S3). However, strong reactivity was also identified at the N-terminus of the Membrane protein undetected in some studies: an extra-virion domain captured by peptide “M1”. Our predicted optimal peptide for M1 comprised the first 19 amino acids of the protein, which protrude in the Membrane protein dimer alongside a short loop between the second and third transmembrane domains on the external virion surface (Fig. 2C). We were struck by an unusual isotype response to M1: anti-M1 IgM was a statistically significant outlier (p<1×10^-7^), with absolutely and relatively higher IgM than other antibody isotypes (Fig. 2D and Fig. S4). Studies that did not profile IgM may have missed this epitope for this reason. M1 showed an unusual pattern where IgM>>IgA>IgG relative to the other epitopes. We examined variants of this peptide to investigate why some studies that had profiled IgM did not identify the reactivity and found that inclusion of even a single extra residue, the tryptophan (W20), diminished the stability of protein substantially, but preserved antigenicity, whereas inclusion of the 5 additional hydrophobic residues (NLVIG) to make a 25-mer, completely abrogated antibody recognition (Fig. S5). This demonstrates that it is not necessarily the case that longer peptides capture more antibodies and structure-agnostic tiling approaches can miss epitopes.

**Fig. 2.**
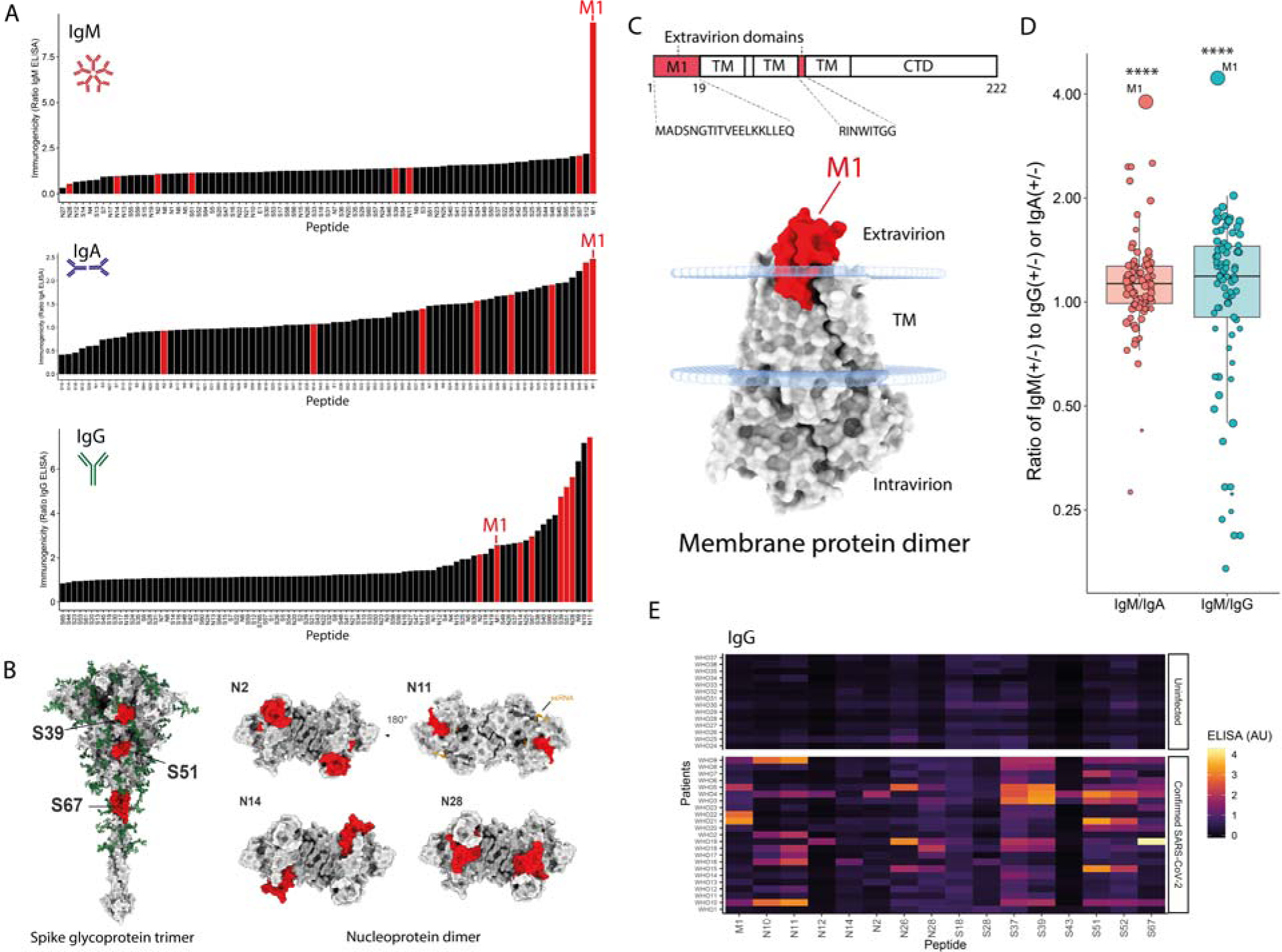
Experimental validation of immunogenicity: an exposed Membrane epitope (M1) is an outlier with an IgM dominant response. **(A)** Selection (red) of eight non-overlapping peptides based on immunogenicity. The ratios of the mean of at least three technical replicates for pooled positive sera to pooled negative sera (representative results of at least two biological repeats (i.e. antigens expressed and purified independently) against individual peptides from the SARS-CoV-2 structural proteins. (B) Position of the selected (red) epitopes on the Spike trimer and Nucleoprotein dimer models. (C) Position of the M1 epitope on the extravirion surface of the Membrane protein dimer (TM = transmembrane domain). (D) Ratio of IgM/IgA (M1 outlier, p=2.1×10^-8^) and IgM/IgG (M1 outlier, p=1.1×10^-8^) for all peptides. Larger size corresponds to higher absolute IgM immunogenicity. P values from Grubb’s outlier test. (E) Heatmap of individual NIBSC reference sera (rows) against peptides (columns) for a selection of epitopes. NIBSC reference panel uninfected individuals (top panel) and individuals after confirmed SARS-CoV-2 infection (bottom panel). Demonstrating individual heterogeneity in the immune response to the eight selected peptides and other peptides from across the structural proteins of the virus.

The specificity of the selected peptides was confirmed using a reference set of 23 PCR-verified SARS-CoV-2 infected and 14 uninfected individuals (NIBSC). In contrast to the aggregate results for whole viral proteins assayed with these sera (Fig. S6), substantial inter-individual heterogeneity in the response resolved to level of epitopes was observed (Fig. 2E). However, single peptides and combinations were able to discriminate unambiguously between infected and naïve individuals (AUC of the ROC for combination of IgG = 0.99, Fig. S7).

### The M1 response is IgM dominant, public, and predictive of known correlates of protection and severity

For testing scarce patient samples, the most immunogenic peptides determined by the ratio of reactivity in pooled positive versus pooled negative (pre-2019) sera were prioritized (Fig. 2A). The responses to these were profiled in sera from patients in independent clinical cohorts recruited in the first pandemic wave in the UK: March-May 2020 (patient characteristics in tables S1-6). The proportions of individuals with detectable IgG responses to each antigen was similar across the cohorts: >80% of individuals in each cohort showed IgG reactivity to at least one of the three spike peptides and >90% had IgG reactivity to at least one of the eight epitopes across S,N, and M (Fig. S8).

Given the apparent dominance of M1 IgM in pooled positive sera, we looked at responses across a second independent cohort of 30 European individuals collected prior to 2019 to verify that these were genuine anti-SARS-CoV-2 reactivity and to determine whether these were antigen-provoked or occured as natural IgM to this target. Consistent with the reference sera (Fig. 2E), no M1 reactivity was observed in any of 30 pre-COVID individuals (Fig. 3A). To determine the antibody kinetics in the convalescent period, random intercept mixed effects models were fitted to longitudinal follow-up samples (Fig. 3B). IgM titres to M1 waned in the 3 months post infection, further consistent with the response having been provoked by infection.

**Fig. 3.**
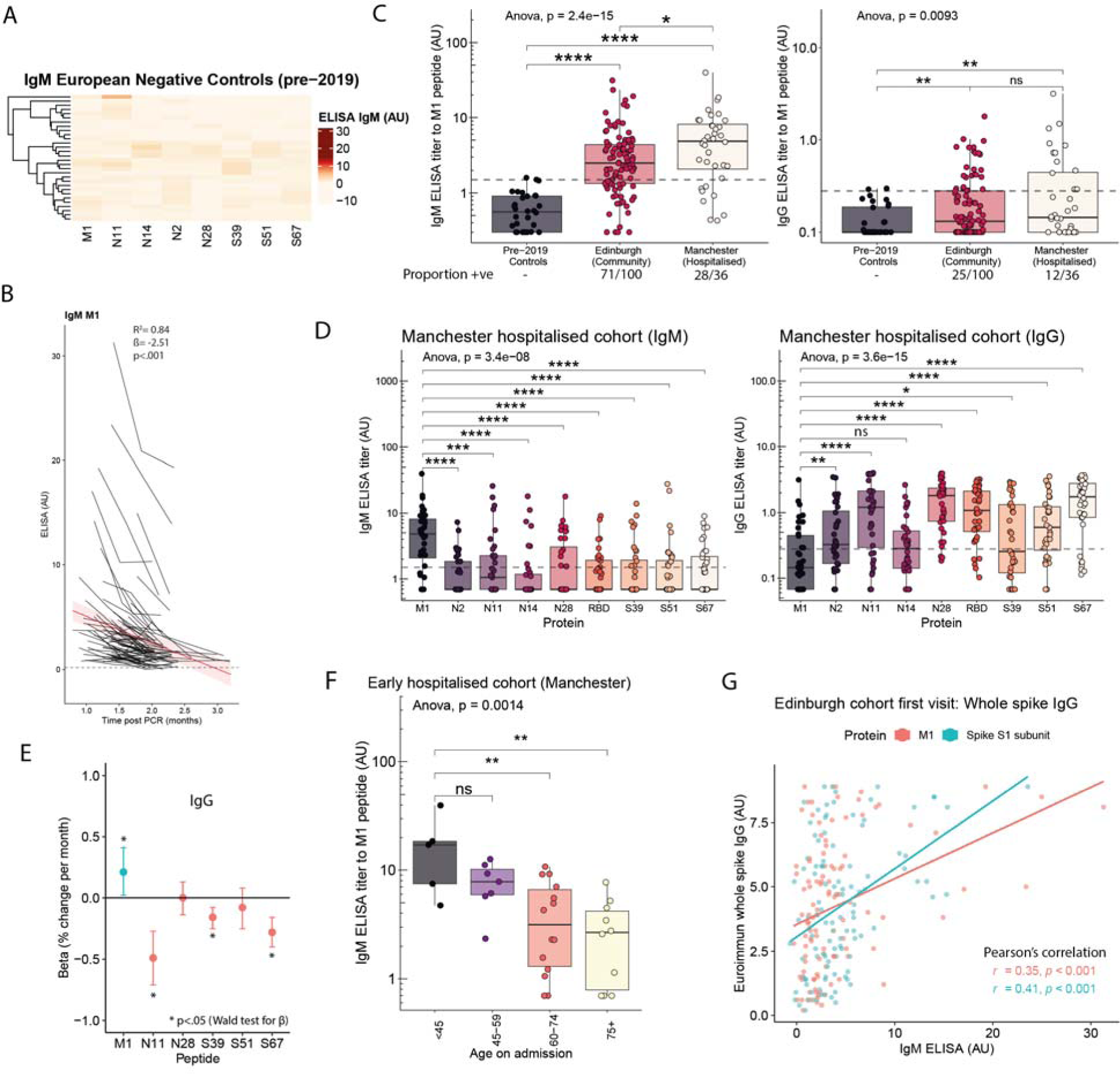
The antibody kinetics for M1 antigen show it provokes a strong IgM response in most exposed persons but isotype class switching to IgG occurs late in only a minority of individuals. **(A)** Heatmap of IgM response by ELISA for 30 European individuals collected pre-2019. (B) Proportion of three NHS clinical cohorts with M1 IgM above the mean + 3SD of the responses in A. (C) IgM and IgG titre to M1 in two clinical cohorts (Edinburgh and Manchester). Dashed line mean + 2SDs of responses for European negative controls (as in A). (D) IgM for the M1 peptide titre tends to be higher than any of the other peptides or the whole receptor binding domain early in the course of infection. (E) Coefficients for time post PCR of random intercept models by antigen. M1 is the only epitope to show a significantly increasing titre in IgG over the three months post PCR. (F) IgM titres to M1 fall over the 3 months post infection and fall fastest for those with highest titres. (G) IgM to M1 and spike S1’ subunit predict aggregate whole spike IgG titre measured by a Euroimmun assay. ns = not significant, * = p<.05; ** p<.01; *** p<.001; **** p<.0001, all post-ANOVA pairwise comparisons are two-sided t-tests.

Most infected individuals in each cohort had detectable M1 IgM responses (Edinburgh: 71/100, Manchester: 28/36) but lacked IgG responses (Fig. 3C). The inverse pattern observed for the other epitopes (Fig. 3D). The IgG response measured for most of the epitopes was stable (N28, S51) or waned (N11, S39, S67) over the 1-3 months post infection. The M1 epitope was again an exception. In the minority of individuals where M1 IgG responses could be detected, they increased significantly from low or undetectable initial levels suggesting ongoing class switch recombination, clonal expansion, or antibody secreting cell differentiation or activation after the acute phase of the illness specific to cells recognizing this epitope (Fig. 3E and Fig. S9).

Comparison of the Edinburgh cohort (of whom only 5/111 were hospitalised) to the Manchester cohort (where all the individuals were hospitalised, required supplemental oxygen, and 27/33 had bilateral chest radiograph opacification), showed a significant tendency for higher M1 IgM in the hospitalised cohort (Fig. 3C). However, differences in the timing of recruitment between these cohorts complicate interpretation of this finding (Tables S1 and S5). Amongst hospitalised individuals recruited early in the course of their infection, there was a strong association with lower M1 IgM titres in older individuals (Spearman’s Rho -0.63, p<.001) (Fig. 3F) which was not evident in individuals sampled at later time points (Fig. S11).

Intriguingly, M1 IgM was found to strongly predict antibody responses to known correlates of immunological protection and severe disease. M1 IgM predicted both whole Spike IgG titre by a commercial assay (Fig. 3G) and pseudovirus neutralisation titre (Fig. S12). The correlation was driven by those individuals with high M1 IgM invariably having high total Spike antibody/pseudoneutralisation, but the inverse was not generally true. This observation was replicated in an independent cohort, finding that M1 IgM predicted whole Spike IgG in blood donors even 2-6 months after infection (Fig. S12). IgM for no peptide other than M1 (including Spike peptides), was significantly associated with whole Spike IgG. IgM for the whole (multi-epitope containing) S1’ subunit of the Spike was correlated with Spike IgG titres and pseudovirus neutralisation, as expected (Fig. S12). Unexpectedly, however, the response to the 19 amino acid M1 was of similar predictive value as the response to the 672 amino acid S1’ Spike subunit, despite the latter containing the ACE-2 receptor binding domain and the N-terminus domain, the sites of most neutralising antibody binding (*19, 20*)
.

### M1 IgM is strongly associated with severe/critical COVID-19 and longCOVID

Motivated by these findings, we investigated whether these antibody responses were associated with clinical outcomes. We tested IgG, IgA, and IgM antibody responses to the eight immunodominant structural epitopes (including M1) and to the whole RBD (a gift from F. Krammer) in another independent cohort of individuals recruited in Oxford, UK. These individuals were recruited either because they had been asymptomatic/mild (non-hospitalised, n=45) or had suffered severe/critical infection requiring ITU admission (n=25), two individuals lacked clinical metadata. Consistent with the Edinburgh and Manchester cohorts, the majority (44/72) individuals had detectable M1 IgM (Fig. 4A).

**Fig. 4.**
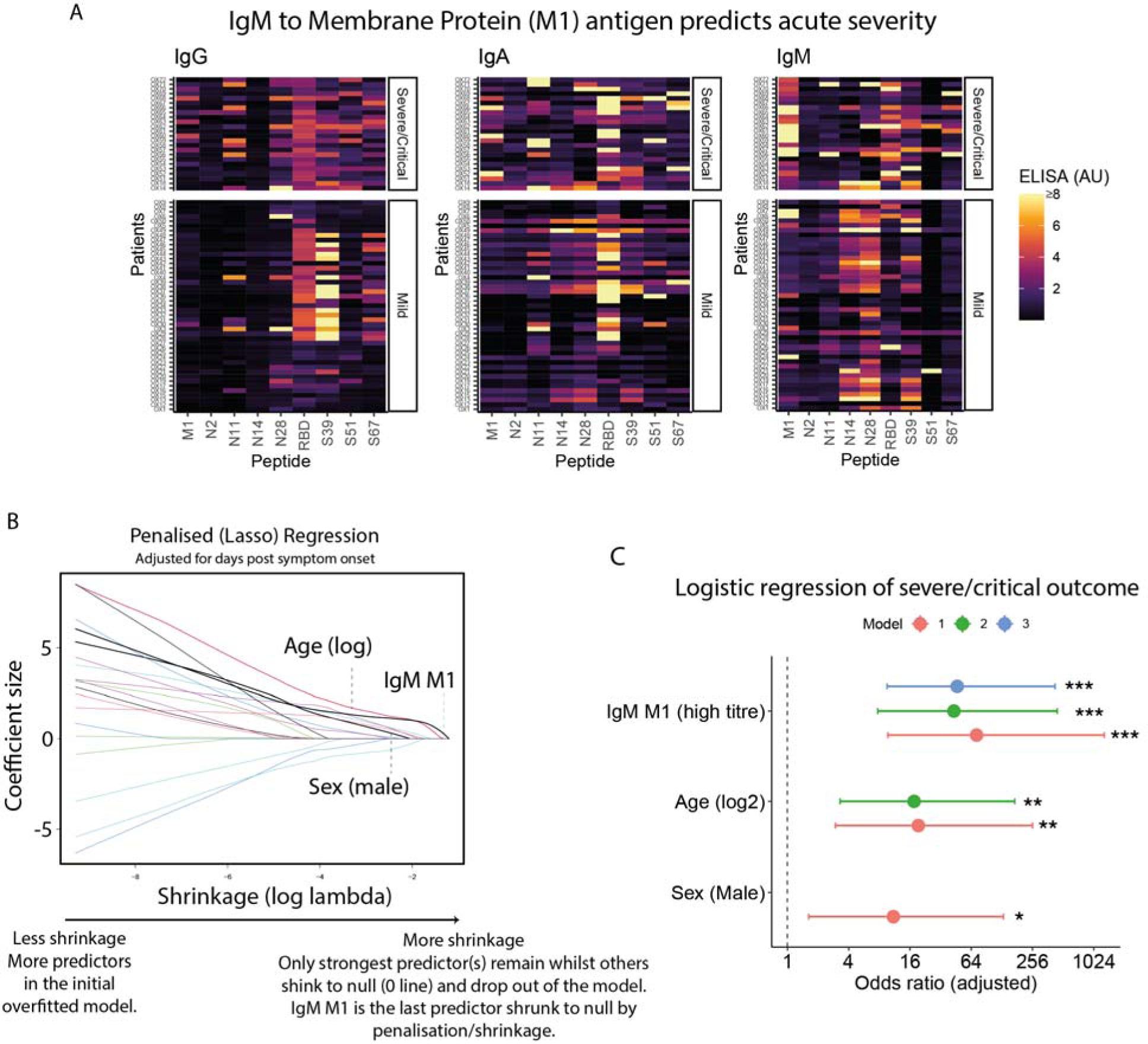
M1 IgM is a strong predictor of severe/critical acute COVID-19. (**A**) Oxford cohort heatmap of individual sera (rows) against peptides (columns) for three immunoglobulin isotypes (column panels). Individuals represented by rows and separated by clinical severity of COVID-19 infection (row panels). (B) Penalized (Lasso) logistic regression demonstrates M1 IgM is the strongest predictor of the outcome (severe/critical COVID-19) of the ELISA responses to the eight peptides and receptor binding domain and age and sex: requires the strongest shrinkage/penalization factor to reduce the coefficient to null demonstrated as it reaches the null (0) coefficient line furthest along X axis. M1 IgM is the only antibody response that is retained in the model as a stronger predictor of severity than age. (C) Multi-variable logistic regression models (without penalization). Points are effect size estimate (adjusted odds ratio) and whiskers 95% confidence intervals. All models adjusted for days post PCR positive. Significance indicated by * p<.05; ** p<.01; *** p<.001 for Wald test for coefficient.

To identify the most important epitope-isotype combinations discriminating severity of disease and to place these in the context of known predictors of disease severity, penalised logistic regression (Lasso) was used. The starting multivariable model includes the logarithm of participant age (to capture the known log-linear association with severity), sex (as males are known to be more susceptible to severe disease) and each isotype for each peptide antigen and the whole RBD (a known immunological correlate of severity). The Lasso algorithm applies an increasingly strong penalty shrinkage factor to multivariable regression coefficients such that they sequentially drop out of the model until eventually only the most predictive variable is left: providing a hypothesis-independent method for ranking variables by predictivity. M1 IgM, was the only antibody response that was a better predictor than age and was a stronger predictor of clinical severity than any other antibody response of any isotype including to the whole RBD (Fig. 4B), and was the last predictor to be shrunk to null by the penalisation. In addition to being correlated with immunological features of severe COVID in independent cohorts, M1 IgM is strongly predictive of clinical outcome.

Next, to estimate the effect size of high M1 IgM, and to examine the statistical interactions with other predictors (age and male sex), multivariable logistic regression models were fitted without penalization (Fig. 4C and Table S3). This reveals that M1 is statistically independent of the effects of age and sex. The point estimate on the effect size was strikingly large: adjusting for

Clinical recovery time post-SARS-CoV-2 infection is variable, even for mild cases. Therefore, given the unusual antibody trajectories and associations with clinical outcome and immunological markers of severe acute disease (Fig. 3E), we were also interested to determine whether there were individuals for whom M1 IgM responses persisted. To explore this, we tested sera from a large cohort of convalescent individuals who had donated plasma to the Scottish National Blood Transfusion service in the 2-6 months after infection. A subset of individuals (22/200) showed persistent M1 IgM (Fig. 5A). K-means (k=5) clustering identified three clusters of individuals with high or medium M1 IgM, one large cluster with relatively low IgM to all the peptides, and a small cluster with IgM to S51 and N peptides (Fig. 5A).

**Fig. 5.**
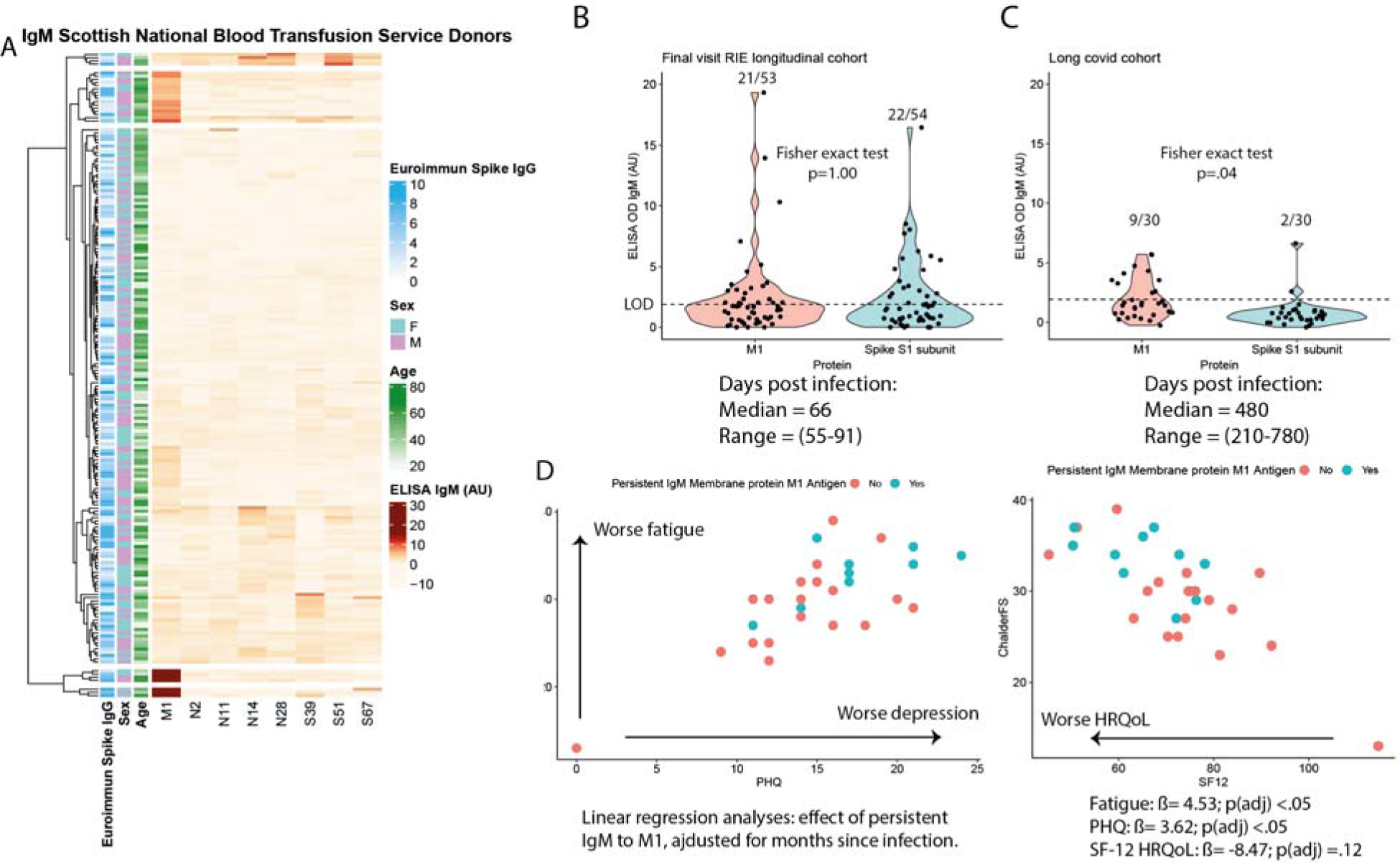
Persistent M1 IgM is associated with longCOVID and symptom burden. (**A**) Heatmap of Scottish National Blood Transfusion plasma donors who donated plasma after infection early in the coronavirus pandemic for trials of therapy with convalescent plasma. K-means clustering identifies two clusters with persistent very high IgM M1 (bottom and second bottom), one cluster with little persistent IgM (large middle group), one cluster with medium IgM M1 (second top), and one cluster with persistent S51 IgM and two nucleoproteins. (B) IgM responses in the final visit of the Edinburgh cohort to M1 antigen and a whole spike S1’ subunit. The whole spike S1’ subunit used rather than other epitopes because no single epitope is close to the M1 epitope for IgM publicness and spike S1’ subunit likely reflects an aggregate of 100s of potential epitopes. LOD = limit of detection for calling positivity based on results in negative control subjects. (C) IgM responses in the long covid cohort to M1 antigen and a whole spike S1 subunit. (D) Chalder fatigue scale (y axis) against PHQ score for anxiety and depression. Subjects represented by points with persistent M1 IgM (cyan) and undetectable M1 IgM (red). (E) Chalder fatigue scale (y axis) against SF-12 score for a health-related quality of life score. SF-12 is comprised of two sub scores with mean 50 reflecting physical and mental domains of quality of life. Here these domains have been combined. Coloured as in D.

Next, we compared IgM titres to the 672 amino acid S1’ Spike subunit and the 19 amino acid M1 peptide in the final visit of the Edinburgh longitudinal cohort and found that 21/53 individuals were still positive for M1 IgM at a median of 66 days post infection and an almost identical proportion (22/54) individuals were still positive for IgM to the much larger multi-epitope containing Spike subunit (p=1.00, Fig. 5B). However, in contrast, repeating this analysis for a cohort of 30 individuals who had been referred by a healthcare provider to a longCOVID study due to persistent cognitive symptoms (> 3 months) post infection (Table S8), we found that 9/30 individuals were persistently positive for M1 IgM, whereas only 2/30 individuals were positive for IgM to the S1’ subunit (p=.04, Fig. 5B). The median time since infection was substantially longer in the long COVID cohort than in any of our other cohorts (480 days). Those with persistent M1 IgM ranged from 240-780 days post infection at the time of sample draw.

In contrast to acute COVID-19, which is associated with a stereotypical clinical syndrome, long COVID is a less well-defined clinical entity where persistent symptoms are probably caused by a variety of mechanisms (*21*). Amongst those who have persistent symptoms will be those with permanent organ damage from the acute illness (*21–23*), and individuals with non-specific symptoms due to other physical or mental illness with onset temporally coinciding with SARS-CoV-2 infection but mechanistically unrelated to immune pathology. Despite the expected heterogeneity, and the small sample size, we found that persistent M1 IgM was associated with a significant 3.62 point worsening on the PHQ-15 score of anxiety and depression (p=0.048) and 4.53 point worsening on the Chalder fatigue scale (p=0.027) (Fig. 5C and Fig. S19). There was also a non-significant tendency for a worse score (-8.47) on the SF-12 health related quality of life questionnaire which combines both physical and mental domains (p=.12) (Fig. S20).

### M1 position on the virion and antibody kinetics suggest it is a T independent antigen

The Membrane protein is the most abundant SARS-CoV-2 protein and forms a homodimer that binds the three other virus structural proteins (S, E and N) (*24–26*). Published analyses of coronavirus virion structure show that the number of Spike trimers per virion ranges between 0 and 50, with 10-20% of virions having no Spikes (*27*). In contrast, each virion comprises ∼2,200 Membrane proteins with M1 exposed in a highly repetitive arrangement at the surface of each virion (Fig. 6A) (*26*). Using cryo-electron microscopy and tomography, Neuman et al. report that M dimers tend to be tightly positioned within coronavirus virions and that M establishes the virion’s spherical shape (*26*). Membrane dimers are topologically arranged as a rhomboids in a plane with sides of each rhomboid approximately 4-5nm, such that the distance across the rhomboid is approximately 7.5nm in one axis and 3.8nm in the other (Fig. 6B) (*26*).

**Fig. 6.**
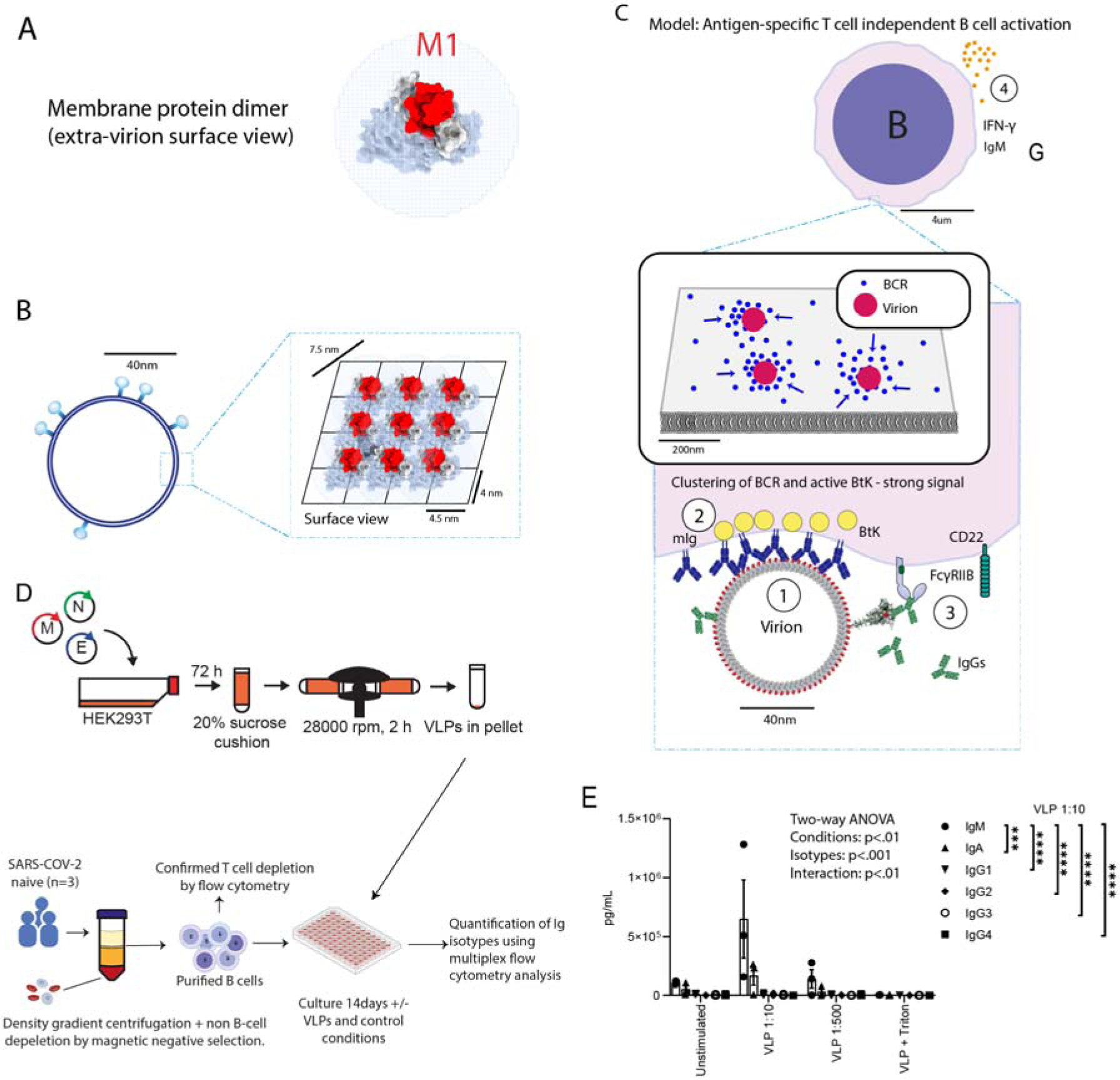
Lack of isotype class switching may be due to T independent B cell activation triggered by the repetitive arrangement of M1 antigen on virions. (**A**) Top down view of the membrane protein dimer from the extra-virion perspective. (B) Repetitive surface of virions with membrane proteins arranged presenting M1 on their surface. (C) Model of T independent B cell activation by repetitive antigen. 1 –arrangement of repeating M1 antigens on a virion clusters membrane bound immunoglobulin (B cell receptors) on the surface of M1 specific B cells. 2 – Cross-linking activation of 10-20 clustered B cell receptors and active BtK is sufficient to trigger calcium influx resulting in B cell activation without T cell help. 3 – secreted antibody (IgG shown) binding to surface epitopes can negatively regulate TI responses by competing with mIg binding or by triggering signaling through surface receptors on the B cell. 4 – Cytokine/IFN release may potentiate or be triggered by TI activation or necessary from other cell types (e.g. Macrophages) and antigen specific IgM is secreted with limited isotype class switching. (D) Schematic illustrating production of spikeless virus-like particles and B cell in vitro stimulation assay. (E) Incubation of B cells with VLPs stimulates IgM secretion compared to unstimulated (PBS) or incubation with disassembled VLPs pre-treated with Triton-X-100, which have otherwise identical protein composition. *** = adjusted p<.001; **** = adjusted p<.0001 for Tukey’s post-hoc pairwise multiple comparison test. Shown are comparisons between IgM and other isotypes for VLPs at high concentration (1:10), all other pairwise comparisons were not significant.

In light of these studies, we hypothesized that the atypical antibody kinetics of the M1 epitope were due to it acting as a T independent B cell antigen (Fig. S13). Antigen-specific T independent (TI) B cell activation occurs in response to repetitive antigens (e.g. polysaccharides or viral capsid proteins) and requires antigenic repeats, scaffolded between 5 and 10nm apart, to recruit a local cluster (10-20) of the ∼10^5^ total B cell receptors on a cell to generate the necessary local signal strength on the B cell membrane (*28*). The abundance and repeating quaternary organization of the Membrane protein in the viral envelope, could provide this signaling in B cells with mIg recognizing M1 (Fig. 6C). TI B cell activation is associated with extrafollicular plasmablast differentiation and the secretion of IgM with isotype class switching limited to specific isotypes. A recent deep repertoire sequencing study of human B cells in response to the T-independent polysaccharide Pneumovax vaccine in humans demonstrated a similar IgM>>IgA>IgG pattern that we observe arising against M1 (*29*).

To facilitate a preliminary test of this hypothesis of TI B cell activation, we produced and purified non-infectious spike-less SARS-CoV-2 virus like particles (VLPs) in cultured human cells (HEK293-T), using methods described previously (Fig. 6D) (*24, 25*)
. The expression of the M, N, and E proteins is sufficient to form Spikeless VLPs (*24, 25*)
. We validated production and purification of VLPs using a protease protection assay and performed transmission electron microscopy (TEM) to confirm the expected morphology (Fig. S14 and Fig. S15).

Next, PBMCs were extracted by density gradient centrifugation from blood drawn from healthy volunteers (n=3) (Tables S7), and untouched B cells were enriched by negative selection, removing T and other non-B cells (Fig. 6D). After culturing with VLPs/controls for 14 days, we found that B cells from 2/3 individuals had secreted IgM, consistent with our hypothesis. This was not observed in unstimulated B cells or B cells co-cultured with VLPs that had first been disassembled by detergent treatment but otherwise were expected to have identical protein composition (Fig. 6E). The pattern of IgM>>IgA>IgG was observed only with VLP stimulation and did not occur in positive controls which were stimulated with R848 (a sythetic agonist of TLR7 and 8) or unmethylated CpG dsDNA (a non-specific B cell activator) (Fig. S16).

## Discussion

Numerous approaches exist for screening linear B cell epitopes, however, as library generation typically tiles k-mer peptides of fixed size with arbitrary overlaps, optimal biophysically stable epitope-containing peptides are usually not synthesized by chance (*16, 18*)
. Which k-mers adopt similar confirmations in isolation as on the whole protein scaffold is also generally unknown. These structure-agnostic methods have low hit rates that limit downstream assays to high-throughput approaches which have important limitations and we find that ignoring structural stability risks missing potentially discoverable and functionally important epitopes (*16, 37*)
. Such high-throughput systems include peptide arrays, or techniques that allow for physical linkage of the binding phenotype to genotype such as bacteriophage or yeast where epitopes are displayed on structures more than 100-fold larger than the peptides themselves (*16, 37*)
. Peptides that express poorly, oligomerize the large display vehicles or aggregate with other peptides can be diminished or enriched, making quantitative interpretation challenging. Aggregation can be particularly an issue for techniques relying on pull-downs of multivalent antibody isotypes (e.g. decavalent IgM), where antibodies and not just the peptides can aggregate and precipitate the display vehicles in complexes. Pull-down of IgM is also challenging as it does not bind Protein A/G. Notably, published studies using these display methods to profile the antibody response to SARS-CoV-2 at the epitope level have tended to omit testing IgM (*16, 37*)
.

The method we describe greatly improves the efficiency of functionally important epitope discovery by incorporating information about the structural stability of the peptides and conformational similarity at the outset. The hit rate is sufficiently high that these predictions can be experimentally validated in diverse downstream applications including low-, medium- or high-throughput techniques. We believe this method has a wide range of possible applications, including in the design of peptide vaccines.

Using the ΔASAr predictions and our screening pipeline, we have identified a Membrane protein epitope with clinically and immunologically important IgM-specific correlates. Despite the vast literature on SARS-CoV-2 serology, to our knowledge, only three relatively small peptide microarray studies have attempted to resolve the IgM response to specific epitopes on the SARS-CoV-2 Membrane protein each within a single clinical cohort (*18, 38, 39*)
, and only one looked at correlates of severity (*39*). Wang et al. profiled responses across the SARS-CoV-2 proteome using an array with 15-mer and 25-mers peptides and found IgM reactivity in only one of eight individuals for the M protein N terminus 15-mer, and no reactivity to the 25-mer peptide, which includes six additional amino-acids not included in our M1 (*18*). Our method predicted that the Wang et al 25-mer, which includes hydrophobic residues from the transmembrane domain, would be the worst of these peptides (highest ΔASAr), and consistent with this prediction, when tested using our pipeline we found the 25-mer to have the lowest reactivity, below the limit of detection for most individuals (Sup Fig. S4). In contrast, and consistent with our findings, Jörrißen et al. tiled 20-mers and found a high proportion (71.9%) of 32 subjects with IgM responses to a peptide which is similar to our M1 (19-mer), and similarly found a high IgM:IgG at two time points after infection (*38*). Hotop et al. used 15-mer peptides, with 10-mer overlaps and identified IgM reactivity to at least one of three peptides comprising the amino acids 1-15, 6-interpret by the concatenation of the C terminus of the E protein to the N terminus of the M protein, a potentially misleading but common strategy in library design which creates chimeric peptides in the array (*31*). These studies illustrate that apparently subtle differences in peptide k-mer design should be expected to affect results and demonstrate the value of accounting for structural similarity (ΔASAr) at the outset (Figs. S2-S4).

Based on the abundance and repetitive quaternary arrangement of the membrane protein within the virion, and the observed pattern of IgM>>IgA>IgG, we hypothesized M1 to be a TI antigen capable of causing extra-follicular B cell activation (*28, 40*)
. B cells are more efficiently activated by membrane bound antigen, as on immune complexes on the surface of follicular dendritic cells, or by repetitively scaffolded antigen on polysaccharides or viral capsids, than by antigen in soluble form. Clustering of multiple activated B cell receptors by scaffolded antigen is thought to provide local amplification of BCR signal strength (*41*). Functionally important antigen-specific TI responses have been previously described for viral capsid glycoproteins antigens including vesicular stomatitis virus (*42*) and polyoma viruses (*43*) but, to our knowledge, not for coronaviruses and not for viral membrane proteins.

Our in vitro data suggest that B cells (depleted of T and other immune cells) can indeed be activated ex-vivo by intact VLPs lacking the Spike protein, where M1 is the only exposed epitope. This supports the hypothesis that the M1 IgM correlations reflect a novel host:pathogen interaction and the identification of a TI antigen within SARS-CoV-2 proteome. However, TI responses are complex and in vivo occur in the specialized architecture of secondary lymphoid tissue and depend on input from cells of the innate immune system (e.g. Macrophages, NK cells and Neutrophils) either directly or by providing supportive cytokines, and on other important situational co-stimulatory signals for example via toll like receptors (*44–46*). There are also considerable differences between the B cell subsets involved in TI responses between rodents and humans, complicating the development of animal models (*29*), and so further work is necessary to understand the mechanisms of this response in detail.

That this host:pathogen interaction may be clinically significant is suggested by our correlative findings in acute and longCOVID cohorts. Our study is predominantly observational, and so we are unable to discriminate causal from non-causal associations. However, the strength of the correlation and statistical independence to age- and sex-is remarkable: if high M1 IgM arose simply as a consequence of severe COVID-19, we would expect it to positively correlate with known risk factors that are themselves predictors of severe disease like age and sex. In fact, we show that M1 is anti-correlated with age in the early days after a positive test in persons who become seriously ill, and at later time points is an independent predictor uncorrelated with these risk factors. Further, it is particularly striking that high M1 IgM is observed in almost all severe cases in these cohorts. Other genetic or immunological correlates of severe COVID-19 have tended to be rare when strongly associated, or weakly associated when frequently observed, which is not the case here. For example, Bastard et al, have described rare (present in <10% of critical individuals) functional auto-antibodies that neutralise 10ng/ml of both INF-α2 and IFN-ω at a 1:10 plasma dilution to be associated with an OR of 67 (95% CI: 4–1109): i.e. strongly predicting severe outcome (*47*). It is notable that the effect sizes are similar: the adjusted OR is 72 (95% CI: 9-1300) for a high M1 IgM in our study.

Our findings of persistent M1 IgM in longCOVID may be worthy of further study as biomarkers of longCOVID are urgently needed to help stratify patients and to objectively score the outcome of trials of therapy and to track the natural history. M1 IgM persistence was significantly associated with fatigue and anxiety/depression, symptoms which place a large burden on quality of life (*21, 23, 51*)
. The biology of fatigue in autoimmune and post-viral conditions is poorly understood, despite the importance to patients. Identifying an immunological correlate of both the presence and degree of symptoms is therefore encouraging. However, many of the participants in the long COVID cohort had identifiable and treatable clinical syndromes contributing to their symptom burden (e.g. depression and migraine) and whilst our results may be consistent with an immunological perturbance – immunological mechanisms have been hypothesized to underlie the aetiology of many such syndromes – more work will be necessary to determine the full clinical significance, if any. The present study was primarily observational and non-hypothesis driven, future studies are ongoing to test the usefulness of this biomarker with predetermined protocols and analyses.

Surprisingly, despite SARS-CoV-2 antigenic evolution, we found that none of the variants of interest or concern have acquired mutations in the prioritized S or N epitopes, which were derived from the original viral reference from Wuhan and only the recent Omicron variants have acquired mutations within the M1 epitope. M:Q19E is common to all omicron subvariants and subvariant specific mutations at the D3 position (D3G, D3N, D3Y,and D3H, Fig. S7) are suggestive of selection pressures at this residue. The M:Q19E mutation has been shown to be associated with increased fitness through an unknown mechanism (*52*). The effect of the M:D3 polymorphism is unknown. The M protein is known to be under particularly strong purifying selection within the SARS-CoV-2 proteome, consistent with its important functions in virion formation. Therefore, the absence of variants with mutations at other positions in M does not imply a lack of function of anti-M1 antibody binding. Similarly, the lack of mutations in the S and N peptides, could be consistent with the virus paying a high fitness penalty for mutations in these structurally stable domains (Fig. S19). The M protein of other coronaviruses is a well-studied model of N- and O-linked glycosylation and strain-specific variation in M glycosylation is thought to determine organ-specific tropism and pathogenicity of murine hepatitis coronavirus by incompletely understood mechanisms (*53–55*). Of note, our screening platform used bacterially expressed peptides which are expected to lack glycosylation and so we may miss some M1-binding antibodies that are specific for glycosylated M1 or M1 only in its homodimer. Our VLP experiments suggest that M is incorporated into virions in both glycosylated and non-glycosylated form. As recent animal work has suggested that the change in disease severity associated with Omicron is not explained by Spike mutations (*57*), M1 mutations may be worth investigating directly for intrinsic differences in disease phenotype in emerging variants, particularly where they impact glycosylation.

The mechanistic triggers that lead SARS-CoV-2, and only certain other coronaviruses, to provoke tissue-specific and temporal uncoupling of inflammation from viral load (*58–60*); breakdowns in immune tolerance; and massive B cell/plasmablast extrafollicular expansion are unknown. The cytokine and immunological response associated with TI antigens overlap with the biomarkers of COVID-19 specific immunopathology (particularly high IFN-gamma, GM-CSF, soluble IgM and increases in plasmablasts) (*28, 61–66*). TI activation of B cells creates expansion of B cells without the oversight of T follicular regulatory cells which have an important role in limiting autoreactivity in B cells expanded in germinal centres. TI B cell activation by viruses is also known to trigger polyclonal TD extrafollicular activation in other B cells, a feature that has recently been shown to be an important characteristic of severe COVID-19 (*36, 67*)
. Finally, the N termini of SARS-CoV and SARS-CoV-2 Membrane proteins show high homology (Fig. S15), in contrast to circulating seasonal human coronaviruses (HCoVOC43, HCoV-229E, HCoV-NL63, and HCoV-HKU1), suggesting that this mechanism may be relevant to other coronaviruses which cause immunopathology. A clear line of sight to therapeutic or preventative intervention would exist if blocking the TI response reduces severity of pathology or inhibits viral replication.

## Methods

### Computational prediction of thermodynamically stable immunogenic peptides

We define ΔASA as the difference between the solvent-accessible surface area (SASA) of the peptide in isolation and the peptide in the context of the full complex, if available, otherwise the monomeric structure is taken. The ΔASA is directly related to the free-energy of hydration of the peptide. To derive ΔASAr, the difference is normalised by the SASA of the peptide:

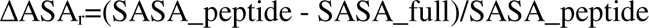

Thus, ΔASAr is a value between 0 and 1, expressing the magnitude to which native contacts of the peptide residues are made with residues outside of the peptide region, rather than being made within the peptide itself. Therefore, a peptide with a lower ΔASAr is more likely to adopt a conformation similar to that in the native structure, which is, intuitively, an important quality of B cell antigens.

For all the peptides occurring in the joint set of SARS-CoV-2 proteins, we generated a peptide-level structural data set by searching the Protein Data Bank (PDB) for SARS-CoV-2 structures from the original “Wuhan” sequence now available at [https://www.uniprot.org/proteomes/UP000464024], as well as closely related structures from SARS-CoV-1 with at least 90% sequence identity to the SARS-CoV-2 proteins. We also included homology models that were available at the time. These include the I-TASSER predicted nucleoprotein model (QHD43423.pdb) made available by the Zhang Lab at https://zhanggroup.org/COVID-19/; the AlphaFold predicted membrane protein monomer modelled as a dimer by the Feig Computational Biophysics Lab, which was made available at https://github.com/feiglab/sars-cov-2-proteins/blob/master/Membrane/M_dimer_new.pdb; and a model of the pentameric envelope protein, which was generated in-house. Briefly, the sequence was submitted to Phyre2 (*68*) on intensive mode on 25/03/2020, followed by sequential alignment in PyMOL to the structure of the SARS-CoV-1 E protein (5x29) and symmetric refinement using GalaxyRefineComplex (*69*).

We calculated the SASA of each peptide in isolation (i.e. without other atoms in the structure file), and in the context of the full complex or the monomeric structure. The SASA was calculated with the software AREAIMOL from the CCP4 suite (*70*), using the default surface probe radius of 1.4. We further limited the data to peptides without missing residues in the structures, and when peptides mapped to multiple structures, we selected the one with the highest resolution.

To generate an initial set of epitope candidates, we ran a wide selection of sequence- and structure-based epitope prediction methodologies to produce linear and discontinuous epitopes for SARS-CoV-2 proteins: BepiPred (*71*), SEPPA (*72*), Discotope (*9*), EPSVR(*73*), and BEpro (also known as PEPITO) (*74*). We also generated a 10-member structural ensemble of the available homology models with the CABS-flex software (*75*), and ran ElliPro (*76*) on each member of the conformational ensemble. This step increases the number of linear epitopes ElliPro can detect, because surface regions have different SASA profiles across the conformers. Then, we searched within immunogenic regions for peptides that have low ΔASAr values. The top scoring peptides for structural proteins were visually inspected in PyMol, resulting in 100 peptide candidates. Peptides for non-structural proteins were selected based on the outputs of the above linear and discontinuous epitope predictors. Initial screening found these to be less commonly B cell epitopes and consequently we focused on Structural epitopes for our studies with limited patient material.

### External validation using publicly available Phage-immunoprecipitation data

To test the utility of the ΔASAr parameter for the prediction of immunogenic peptides, we made use of publically available peptide-tiling experiments with associated immunogenicity scores. Two such data sets were available at the time of our screening experiments (after we had established our synthetic biology pipeline; these were not available at the time that we selected our prioritized peptides), which will be further referred to as VirScan (*16*) and ReScan (*17*). VirScan employed 56 and 20 amino acid (aa) long peptides, while ReScan used 36 aa long peptides in a phage display setting to identify those most enriched for binding to immunoglobulins extracted from human patient sera. In case of the VirScan peptides, we downloaded Table S4 provided by Shrock et al., 2020, and averaged over the IgG Z-scores for all positive and negative cases and took the difference as the immunogenicity signal. In case of the ReScan data, we downloaded the data files from https://github.com/UCSF-Wilson-Lab/sars-cov-2_ReScan_VirScan_complete_analysis, then extracted all SARS-CoV-2 specific peptides’ read counts from the pancorona folder, excluding healthy patient sera and controls. Subsequently, for each peptide we calculated the mean read count, filtered for a minimum of 50 reads, and took these values as the immunogenicity signal.

We then asked whether lower ΔASAr values are associated with a higher immunogenicity signal. To assess this, we leveraged the adjacency of the peptide tiles, i.e. the fact that they overlap and thus cover a large fraction of the same potentially immunogenic region. In the ReScan data, the overlap is exactly 50%, while in the VirScan data 20-mer tiles have a distance of 5 aa between their midpoints (75% coverage), and 56-mer peptides have 28 aa between their midpoints (50% coverage). We calculated in any given peptide pair the number of times the peptide with the lower ΔASAr is the one with the higher immunogenicity score. We show this analysis in Figure S3A with results pooled for 56-mer and 20-mer peptides, filtered at different Z-score cutoffs. The results suggested that the hypothesized relationship holds in the large majority of cases when we only consider peptide pairs covering the most immunogenic regions (16 out of 20 pairs, p = 5.9 × 10-3, binomial test). To approach the problem from a different angle, we binned members of adjacent peptide pairs in the ReScan data set according to “lower” and “higher” ΔASAr values, and compared the distribution of mean read counts between the two bins. This analysis revealed that the peptides with lower ΔASAr have a significantly higher immunogenicity signal than their adjacent partners with higher ΔASAr values (228 pairs, p = 8.6 × 10-3, Wilcoxon signed-rank test). Furthermore, the trend does not hold to a significant extent when the SASA of the peptide (ASApeptide) or the relative accessible surface area of the peptide (RASApeptide), calculated as the ratio of the observed SASA to the sum of the residues’ theoretical maximum, is considered, suggesting that the origin of the trend is not simply that immunogenic regions tend to be more exposed to solvent. These analyses demonstrate that this simple structural measure, ΔASAr, is useful for the prioritisation of computationally generated peptides for laboratory-based immunological screening experiments.

### Protein model visualisations

Structures of SARS-CoV-2 proteins for visualisation in figures were produced as follows. The Feig lab Membrane protein model was used to create a membrane immersed form of the dimer on the PPM 3.0 server (*77*). The fully glycosylated spike protein was acquired from the CHARMM-GUI Archive in the open and closed conformations (6VSB 1_1_1 and 6VXX 1_1_1), which are freely available at https://www.charmm-gui.org/?doc=archive%lib=covid19. The nucleoprotein dimer was generated in-house using a homology modelling approach. A scaffold structure was generated by submission of the full length N-protein sequence to Phyre2 (*68*) on intensive mode on 02/04/2020. The N and C-terminal domains were replaced with the corresponding regions of 6m3m chain A and 2gib chain A, respectively, and a homodimer was created by structural alignment to the 2gib biounit in PyMol. The structure was subsequently refined with the software GalaxyRefineComplex (*69*) using a C2 symmetric refinement protocol. For visualisation in figures, a 10-mer ssRNA was transplanted from the structure 7act. Figures were prepared with UCSF ChimeraX v1.3 (*78*), PyMol (v2.4.0, Schrodinger, LLC), and Adobe Illustrator (2022).

### Synthetic biology peptide-fusion protein pipeline

#### Vector construction

After identification of putative immunogenic peptides, DNA sequences were designed corresponding to the fragments flanked with recognition sites for the type IIS restriction enzyme BsaI. A second, unique restriction site was also included for diagnostic purposes. These were synthesised by a commercial manufacturer (IDT) in the form of eBlocks Unique overhangs, generated by BsaI digest, at the 5’ and 3’ ends of the DNA fragments allowed the directional insertion into expression vectors using Golden Gate cloning. New vectors were made (i) for expression in mammalian cells and (ii) for expression in bacterial cells, and designed to include useful characteristics to enable stable, high level protein expression. For both mammalian and bacterial vectors a similar design framework was used so that DNA libraries could be efficiently ligated into both vectors using a high-throughput robotic platform. The general outlayof the vectors consists of the following components: a host-specific promoter, histidine purification tag, protease cleavage site, fusion protein domain, afragment with two BsaI directional cloning sites, and a termination/polyadenylation site (Fig. S1). The bacterial construct uses an inducible T7-LacO promoter and contains a LacI gene in its backbone. To anchor the small, SARS-CoV2 derived peptides it has a GST fusion protein domain, whilst the mammalian construct uses a rabbit Fc fusion domain. The mammalian vector also contains an IL2 leader sequence for secretion in the culture media. Design considerations and experimental testing of the various vector components to identify those with the best and most consistent protein expression will be described in detail elsewhere. DNA libraries were cloned into the vectors as described; individual clones were confirmed by diagnostic restriction enzyme digest and verified by Sanger sequencing.

#### Bacterial protein expression and robotic affinity chromatography protein purification

35ul of T7 Express E. coli competent cells (NEB) were heat-shock transformed with ∼25ng of plasmid DNA and outgrown in 200ul SOC media for an hour at 37^0^c in 1.5ml Eppendorf tubes shaking at 500rpm on a benchtop heated shaker. 100ul of outgrowth was then added to 4ml Luria-Bertani (LB) media with 100ug/ml ampicillin in 24 deep-well plates, covered with aeraseal and grown overnight at 250rpm, 37^0^C, until late log phase. The following morning these were subcultured and grown to an OD of 0.4-0.6, shifted to 18^0^C and IPTG was added to 0.5 mM. After overnight expression, the bacteria were pelleted by centrifugation at 4000G, media was discarded and pellets washed in PBS. The pellets were kept frozen at -20^0^C until purification.

E. coli pellets were resuspended in 500ul of BugBuster (Millipore, 70584) buffer with 1ul of 400x PMSF and 1ul of Benzonase in the 24 well plates. Resuspended samples were shaken for 30mins at 250rpm in 25^0^C incubator for lysis then centrifuged at 4000G at 4^0^C with this temperature maintained for all subsequent steps. The supernatant of each lysate from four 24 well plates was transferred to a 96 deep well plate for purification in a Kingfisher Flex robot (Thermo Fisher) using a custom program. Samples were added to 100ul Pierce Nickel-NTA magnetic agarose bead suspension (ThermoFisher, 78605) 25ul settled bead volume, after beads were equilibrated in (50mM sodium phosphate, 0.3M NaCl, 0.05% Tween 20, pH 8.0) with 15mM freshly prepared imidazole. After binding samples with mixing for 1 hour, protein-bound beads were magnetically collected and were washed twice in the same buffer with 30mM imidazole. Proteins were then eluted in two fractions from the beads in 100ul of elution buffer (detergent free base buffer with 500mM imidazole). Eluted proteins were immediately desalted using Sephadex columns (PD Multitrap G-25, Cytivia, 28918006) into storage buffer (PBS with 10% glycerol).

#### Mammalian protein expression

Expi293 cells were transfected in 6-well flat-bottom plates using expifectamine transfection kit as per manufacturers recommendations and were grown at 37^0^C in 8% CO2 in expression media on an orbital shaker at 100 RPM. Supernatant was harvested at day five and kept at -20^0^C until purification. The mammalian protein was purified by adding the supernatant directly to 100 ul of magnetic beads. Beads were washed with 1ml base buffer and eluted in 100ul elution buffer. Samples were desalted using Sephadex columns (PD-10 MiniTrap Sepahdex G-25) into storage buffer.

#### SDS-PAGE electrophoresis

Purified fusion-protein samples were prepared for gel electrophoresis with Laemelli loading buffer (final conc. 31.5mM Tris-HCL, pH 6.8, 10% glycerol, 1% SDS, 0.005% Bromophenol Blue) with 100uM dithiothreitol and heated for 5 mins at 90^0^C, and were run out on SDS-PAGE gels (4-12% Bis-His in MOPS buffer). Gels were washed in warm distilled water three times to remove SDS and stained for one hour after being heated once in a microwave with 0.2% w/v Coomassie Brilliant Blue G250 in 0.1M citric acid and then de-stained in distilled water overnight before photographing.

#### Protein Quantification

Protein quantification was performed after desalting samples as described above, as comparison of the reproducibility of various protein quantification methods found all to be unreliable in the presence of imidazole at either 300mM or 500mM. NanoOrange (Invitrogen, N6666) showed acceptably low protein-protein variability and was practical for high throughput quantification using manufacturers protocol. Proteins were quantified at a 10x dilution for mammalian expressed proteins and 200x dilution for bacterial expressed proteins. Duplicates were read in 96-well black OptiPlates at ∼485/590nm.

### Experimental validation of immunogenicity

#### Subjects

In total 605 serum samples were analysed for 454 individuals who had been infected with SARS-CoV-2 and 50 samples from 50 individuals collected prior to 2019. All subjects were recruited following protocols approved by local ethics committees. Informed consent was collected following committee recommendations in all cases. Further processing of patient samples and deidentified personal data was performed according to procedures, risk assessments and protocols under over-arching ethical approval from the Edinburgh Medical School Research Ethics Committee of the University of Edinburgh (reference: 21-EMREC-010). Local ethical approvals and cohort recruitment details for sample collection are detailed under cohort descriptions below.

#### Serum samples for assay development

Peripheral blood samples (serum and plasm) for assay development were obtained after informed consent from volunteers at University of Edinburgh. These were centrifuged immediately after collection and aliquoted. Aliquots of plasma/serum samples were heat-inactivated (56^0^C for 1h) and stored at 4^0^C until use.

#### Technopath pooled and NIBSC reference sera

CE-marked anti-SARS-CoV-2 verification panel of 37 samples was used in ELISAs for confirmation of epitope immunogenicity. Samples contain human plasma and Bronidox at 0.05% (w/vol) as a bacterial growth inhibitor. 23 samples were from known anti-SARS-CoV-2 positive individuals (WHO1-23 in this study) and the remaining 14 reported as SARS-CoV-2 naïve individuals (WHO24-37). No clinical information was available beyond infection status for these samples. Laboratory values were provided for results of commercial antibody tests (Fig. S4).

#### Manchester

*Ethics:* Cohort ethical approval was obtained from the National Research Ethics Service (REC reference 15/NW/0409 for ManARTS and 18/WA/0368 for NCARC).

Subjects were recruited from Manchester University Foundation Trust (MFT), Salford Royal NHS Foundation Trust (SRFT) and Pennine Acute NHS Trust (PAT) under the framework of the Manchester Allergy, Respiratory and Thoracic Surgery (ManARTS) Biobank (study no M2020-88) for MFT or the Northern Care Alliance Research Collection (NCARC) tissue biobank (study no NCA-009). Informed consent was obtained from each patient, clinical information was extracted from written/electronic medical records including demographic data, presenting symptoms, comorbidities, radiographic findings, vital signs, and laboratory data. Patients were included if they tested positive for SARS-CoV-2 by reverse-transcriptase– polymerase-chain-reaction (RT-PCR) on nasopharyngeal/oropharyngeal swabs or sputum during their in-patient admission for COVID-19. Patients with negative nasopharyngeal RTPCR results were also included if there was a high clinical suspicion of COVID-19, the radiological findings supported the diagnosis, and there was no other explanation for symptoms.

Samples (100ul serum) on dry ice and de-identified subject data were transferred to the corresponding authors under a material transfer agreement between the University of Manchester and the University of Edinburgh.

#### Edinburgh cohort

*Ethics:* Cohort samples were obtained under ethical approval granted through the NHS Lothian BioResource (SR1407) and London-Brent Research Ethics Committee (20/HRA/3764 IRAS:28653). All participants gave written and informed consent for serial blood sample collection.

Samples (300ul serum) and deidentified subject data were transferred to the corresponding authors internally withing University of Edinburgh without material transfer agreement.

#### Oxford cohort

*Ethics:* Severe/critical samples were collected under Sepsis Immunomics project (Oxford REC C, reference:19/SC/0296) or ISARIC/WHO Clinical Characterization Protocol for Severe Emerging Infections (Oxford REC C, reference 13/SC/0149) or consenting into the CMORE study protocol (Northwest–Preston REC, reference 20/NW/0235). Asymptomatic/mild samples were collected under the Gastro-intestinal Biobank Study in Oxford: 16/YH/0247, approved by the research ethics committee (REC) at Yorkshire & The Humber -Sheffield Research Ethics Committee on 29 July 2016, which has been amended for the purpose of the COVID-19 substudy on 8 June 2020.

Severe/critically ill patients were recruited from the John Radcliffe Hospital in Oxford, UK, between March and May 2020 by identification of patients hospitalised during the SARS-CoV-2 pandemic. Time between onset of symptoms and sampling was obtained for all patients. Written informed consent was obtained from all patients. All patients were confirmed to have tested positive for SARS-CoV-2 using the reverse transcriptase polymerase chain reaction (RT-PCR) from an upper respiratory tract (nose/throat) swab tested in accredited laboratories. The mild/asymptomatic group were predominantly healthcare workers recruited from Oxford University Hospitals NHS Foundation Trust after a positive SARS-CoV-2 PCR test in April-May 2020.

Blood samples were collected and separated into plasma by centrifugation at 500 g for 10 mins. Plasma was removed from the uppermost layer and stored at −80°C. Samples (60-150ul plasma) on dry ice and deidentified subject data were transferred to the corresponding authors under a material transfer agreement between the University of Oxford and the University of Edinburgh.

#### Scottish National Blood Transfusion Service (SNBTS) cohort

*Ethics:* Use of samples for this study were approved by the SNBTS Research and Sample Governance Committee (reference: SG2021-25).

Scottish blood donors provided informed consent for microbiological testing at their donations under SNBTS Blood Establishment authorization. Samples were collected between 01/10/20 and 21/03/21.

#### Long COVID cohort

*Ethics:* The long COVID study was approved by North of Scotland Research Ethics Committee ref 21/NS/0035.

The long COVID samples were taken from consecutive participants in a cross-sectional prospective cohort study of adult patients with persistent cognitive symptoms that lasted for greater than 3 months after COVID-19 in Edinburgh, UK, referred by hospital clinicians or general partitioners, Patients were reviewed at the Anne Rowling Centre for Regenerative Neurology at the Royal Infirmary of Edinburgh by consultant neuropsychiatrists for detailed clinical phenotyping and plasma was taken at time of consultation. Samples and deidentified subject data were transferred to the corresponding authors internally within University of Edinburgh without material transfer agreement.

#### Indirect ELISA

Flat-well, Greiner Microlon-coated high protein binding 96-well ELISA plates wells were coated in duplicate with 50ul of 1.25ug/ml of recombinant protein suspended in coating buffer (15mM Na2CO3, 35mM NaHCO3, pH 9.3-9.6) and incubated at 4^0^C in a stationary humidified container overnight. After binding, the plates were washed four times with PBS-T (1.9mM NaH2PO4, 8.1mM Na2HPO4, 150mM NaCl, pH 7.2-7.4 + 0.05% Tween-20). Plates were then blocked with blocking buffer (1% skimmed milk powder in PBS-T) for 90 minutes at 37^0^C. After washing, serum samples were diluted 1/50 in blocking buffer before 50ul was added to the desired wells for 90 minutes at 37^0^C. Plates were washed again as before and secondary antibody in blocking buffer was added: rabbit anti-human horse raddish peroxidase anti-IgG (1/3000), -IgA (1/500), or -IgM (1/500) (Dako, Agilent Technologies Denmark). After an hour of incubation, plates were washed and bound secondary antibodies were visualised by adding 100ul per well of 0.04mg/ml O-phenylenediamine dichloride (OPD, Sigma) and 0.012% hydrogen peroxide in development buffer (24.5mM citric acid monohydrate, 52mM Na2HPO4, pH 5.0). The reaction was quenched using 2M H_2_SO_4_ based on A450nm of 0.7-0.9 and optical density (OD) at 492nm. The reaction-quenched plates was then read and recorded on a Labsystems Multiskan Ascent plate reader, plates were photographed, and data imported for analysis using R. Preliminary experiments were performed with dilutions of pooled and individual sera to determine the optimum antigen (1.25ug/ml) and serum (1/50) concentration for the assays. Sample data points were excluded only where during ELISA a pipetting error occurred and was noted, or if controls on the plate were not satisfactory the whole plate was repeated. Samples were used up in the testing of samples and so antigens were prioritized based on limited samples and some samples were exhausted before all antigens could be tested.

#### ELISA analysis

ELISA results were corrected based on the blank controls on each plate and normalized to a positive plate control. The ratio of the sample reading to the positive plate control reading allowed for quantitative interpretation of antibody titre at a single dilution sparing clinical samples. All ELISA “titres” referred to in this study are these adjusted ratios and are expressed in arbitrary units (AU). Dilution experiments with abundant pooled and negative sera ensured that the antigen concentration and serum dilution was in the linear range for all of the epitopes. This was achievable as only one or a small number of epitopes are expected for each peptide and peptides are attached to the same large fusion protein reducing protein-protein variability in quantification. Replication experiments performed by different study investigators on different days ensured the methods gave consistent results. Samples were assayed in triplicate or in duplicate when serum sample scarcity required. The investigator performing the ELISAs was blinded to the clinical status and participant characteristics for all samples.

Ratio of antibody responses used to rank epitope immunogenicity are mean of 3 titre results from pooled positive samples/mean of 3 titre results for pooled negative samples. Positive and negative pools were run simultaneously on the same plates at the same dilutions of primary and secondary antibodies and antigen. Arbitrary interpretation of these immunogenicity ratios is as follows: Positive (Ratio >=1.5), Negative (Ratio <1.2), Borderline (1.2=<Ratio<1.5).

For binary (positive/negative) interpretation of ELISA assays, we took a mean of blank corrected technical replicate ODs before dividing by the mean of positive plate control (whole spike S1’ subunit and pooled positive sera 1/50 dilution). The cut-off for positivity for each antigen was determined for each secondary antibody by the plate-corrected mean of negative controls + 3 standard deviations.

#### Pseudovirus neutralization assays

Pseudovirus neutralization experiments have been previously described for both the Edinburgh cohort and the Oxford cohort. These data were used in the analyses described in this paper (Fig. 5F and Fig. S9).

The Edinburgh cohort pseudovirus experiments/results have been previously described in detail (*79*). A brief summary of those method is as follows: 5x serially diluted serum from convalescent individuals were incubated with SARS-CoV-2 pseudotyped virus for 1 h at 37 °C. The mixture was subsequently added to 293TAce2 cl22 cells at a starting serum dilution of 1:50. Nanoluc Luciferase activity in lysates was measured 48 hours post-inoculation using the Nano-Glo Luciferase Assay System (Promega) with the Glomax Navigator (Promega). Relative luminescence units were normalized to those derived from cells infected with SARS-CoV-2 pseudotyped virus in the absence of serum. NT50 are half-maximal neutralization titers for sera determined by four-parameter nonlinear regression (GraphPad Prism).

### Analysis of experimental results

#### Linear mixed effects regression analyses of Edinburgh longitudinal samples

We fitted linear mixed-effects models (estimated using restricted maximum likelihood) for IgM and IgG antibodies on each protein including a random intercept for the participant. For fitting the models we included only participants who had at least one detectable antibody response of the isotype in question to the epitope in question in the first three study visits. Mixed-effects models were chosen to assess the effect of time post infection whilst accounting for the repeated measurement of participants. For each observation *i*, we modeled the antibody response Yi as:

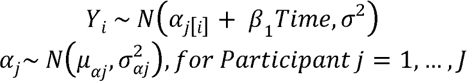

Where β1 is the fixed-effect slope coefficient for *Time* the number of days after the first SARS-Cov-2 PCR+ result was returned that the blood sample was drawn. *α*_j_ represents the participant-level normally distributed random intercept. We examined the total explanatory power (conditional R2) and residual plots to check assumptions (linearity, random distribution of residuals and homoscedasticity) for each model. We also considered models with additional fixed effects for covariates (age, sex, hospitalisation status, and a proxy for overall antibody titre in the Euroimmun IgG assay) and found that the inclusion of these terms did little to improve model fit and did not materially change the conclusions drawn from the modelling about the rate of waning of isotypes for specific epitopes. This suggests that these covariates tend not to greatly effect the rate at which antibodies wane beyond their effects on the starting titre, which is already accounted for in the participant-level random intercept. Models were fit using the lme4 package in R statistical software.

#### Lasso penalized regression

Examining the response for three antibody isotypes (IgG, IgM, IgA) for eight epitopes and the receptor binding domain, and covariates for sex, age, and days since symptom onset, yields 30 parameters for the 70 Oxford cohort participants: i.e. a high ratio of parameters (p) to subjects (n). We therefore used a penalized regression approach to discriminate the most predictive immunogenic epitopes (reducing the p:n). The least absolute shrinkage and selection operator (Lasso) method was adopted as it allows for variable selection during regularization and can be applied to generalized linear models. In a Bayesian interpretation, Lasso regression can be interpreted as linear (or generalized) regression where the coefficients have Laplace prior distributions (peaked at zero) leading to a tendency for coefficients to shrink to zero. This feature is an advantage over other regularization techniques (e.g. Ridge regression) as it aids interpretability by eliminating less-predictive variables from the model. Since we intended to include age, sex, and days post PCR of sampling, as covariates in subsequent models, we were interested to identify the single most predictive immunogenic epitope for effect size estimation in non-penalized regression models, rather than selecting the optimum regularization parameter and multiple isotype-epitope responses. However, we also examined the AIC for sequential values of the regularization parameter to identify the optimum value and performed sensitivity analyses to examine the approach to variable transformations on the results of selection.

The age variable was log transformed, sex was included as a binary variable, and time post PCR result was coded in days. Because we were comparing different secondary antibodies used at different concentrations simultaneously and because we had limited sample volume (60ul – 100ul) with which to test all isotypes for all epitopes and so had used the ratio of a single dilution (1/50) of serum to a known positive plate control (Technopath positive serum to whole spike S1’ subunit) to calculate the arbitrary units of ELISA titre, we interpret these titres as semi-quantitative. The validity of this approach is evidenced by the expected waning in longitudinal responses and the consistency of our results identifying immunodominant antigens with those reported by others. However, titres will not be directly comparable between isotypes due to the issue of different secondary antibodies meaning that these variables required a some form of normalizing transformation. Each sera was used at a dilution of 1/50 and each antigen at 1.25ug/ml which prior experiments had suggested would mean that most subjects with detectable responses were on the linear part of the ELISA response curve, however, those with very high titres will be outwith the upper limit of the linear range of our assay and so discriminating between extreme high titres would be potentially misleading. Therefore, we dichotomized each antigen based on the median response in the whole cohort (n=70) to represent either low (or no) versus high antibody responses. We determined not to include any antibody isotype where <5% of the cohort were responders (which excluded the IgG response to the N2 epitope) to prevent misleadingly incorporating responses below the lower limit of detection of the assay. We performed a secondary analysis including all variables but dichotomizing at the lower of either: 1/ the lower limit of detection of a response for each epitope-isotype combination based on the mean response of the 50 individual negative controls + 3SDs, or 2/ the median response in the cohort. These two approaches we found to give substantially the same result and the simpler median dichotomization is therefore reported in Fig. 5B due to being easier to interpret.

We performed further sensitivity analyses using a cutoff of the 65^th^ percentile (45/70) rather than the median as the cohort were not a population based random sample of persons infected with SARS-CoV-2, rather essentially were a comparison of an arbitrary number of asymptomatic/mild individuals (n=45) to severe/critical (n=25) individuals. This sensitivity analysis corroborated the prior results highlighting the importance of the M1 IgM response. Lasso regression models were fit using the glmnet package in R statistical software.

#### Multivariable logistic regression models

After identifying the M1 IgM as the strongest immunological predictor in Lasso models, we fitted three logistic models (estimated using maximum likelihood) without shrinkage to predict COVID-19 disease severity with age and M1 IgM as predictors (Fig. 5C). As above, we log (base 2) transformed age given the well described log-linear association of age and SARS-CoV-2 infection fatality rate. Exponentiated age coefficients can be interpreted as the odds ratio associated with doubling a participant’s age, adjusting for the other variables in the model. We dichotomized M1 IgM at the lower of the median value or the lower limit of detection of the assay for the responses to each isotype and each epitope, as before for Lasso regression.

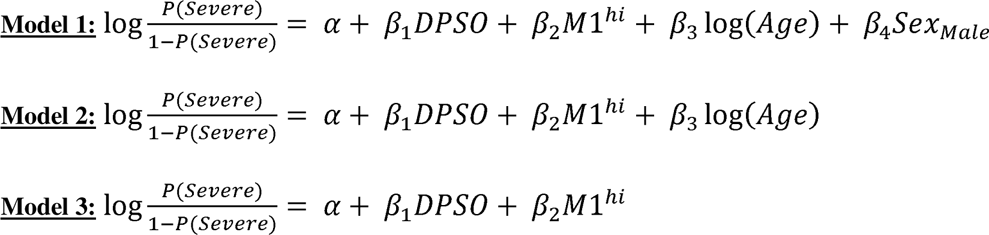

Where β1 is the coefficient for days post symptom onset (*DPSO*), β2 is the coefficient associated with a high M1 IgM (*M1^hi^*), β3 the coefficient for the logarithm (base 2) of age, and β4 the coefficient for male sex (*Sex_male_*). We examined the total explanatory power of all three models using Tjur’s R2 (ref) and inspected residual plots to ensure assumptions of the model (linearity, random distribution of residuals and homoscedasticity) were not violated. We performed sensitivity analyses as for Lasso penalized regression: using a cutoff at the 65^th^ percentile (45/70) rather than the median, and using the untransformed raw ELISA titre and the log transformed ELISA titres. Sensitivity analyses did not substantially improve model fit or change the interpretation of the model, the coefficients for age, sex, and having a high IgM remained statistically significant, positive, and large. Confidence intervals for logistic regression models were calculated using the profile likelihood method.

#### Dimensionality reduction methods

To reduce the dimensionality of the 8 epitopes + RBD, and 3 antibody isotypes, we employed uniform manifold approximation and projection (UMAP) to represent patterns in two-dimensional space. Antibody titres were log transformed and scaled by the standard deviation of the response for each peptide. Similar results were observed using unscaled and untransformed data. UMAP were performed using the UMAP package in R statistical software.

#### Multiple sequence alignments

Multiple sequence alignments were performed using MUSCLE with default parameters. Alignments were manually adjusted where necessary and figures prepared using Snapgene software (Insightful Science), Jalview, Adobe Acrobat and Adobe Illustrator.

#### Analysis of SARS-CoV-2 variant mutation frequency

Sampling of public sequencing data to investigate the prevalence of mutations in M1 epitopes was performed using tools available at www.nexstrain.org using GISAID data (*80*).

#### Statistical software

Statistical analysis were performed in R v4.2.0 and RStudio v1.2.5019. A complete list of packages with version numbers are provided in the supplementary materials.

### Experimental testing of TI B cell activation hypothesis

#### Production and purification of SARS-CoV-2 virus-like particles (VLPs)

HEK293T cells used for VLP production were maintained in DMEM supplemented with 10% FCS, 100μg/μl penicillin, and 100μg/μl streptomycin sulphate. HEK293T cells in T-175 flasks were co-transfected with plasmids for SARS-CoV-2 M gene (18.75μg), E gene (18.75μg)1, and N gene (12.5μg) (gift from Paul Digard) using 100μl Lipofectamine 3000 transfection reagent and 100μl P3000 reagent (Invitrogen, L3000001) according to the manufacturer’s instructions. VLPs were harvested from cell culture supernatant 72 h after transfection based on a previously described method (*81, 82*)
. Briefly, cell culture supernatant was clarified by centrifugation at 1000xg for 10 min, clarified supernatant was transferred to ultracentrifuge tubes and underlaid with a 20% sucrose cushion using a 4” blunt needle. VLPs were pelleted by centrifugation for 2h at 4°C, 28,000rpm using an SW28 Ti swinging bucket rotor. Supernatant was removed and ultracentrifuge tubes inverted for 5 minutes on paper towel to remove residual supernatant. Pelleted VLPs were resuspended in 200μl of PBS per T-175 flask used for VLP production. VLPs were aliquoted, snap frozen, and stored at -80°C until use.

#### Protease protection assay to assess VLP integrity

VLPs were assessed for lipid membrane integrity using a protease protection assay. Equal amounts of VLPs were treated with 1% triton-X-100 or 1X Trypsin 1:250 (Gibco) as indicated and incubated at 37°C for 1h. VLPs were then mixed with 4X SDS-PAGE loading buffer and heated at 95°C for 5 min prior to SDS-PAGE and Western blot analysis.

#### Western Blotting

Proteins were separated by SDS-PAGE and transferred to 0.45μm PVDF membranes. After blocking with 5% skim milk in TBS-T, membranes were incubated with the indicated primary antibodies (0.25μg/ml sheep anti-N or 0.25μg/ml sheep anti-M)1, followed by secondary antibody horseradish peroxidase (HRP)-conjugated rabbit anti-sheep IgG (Santa Cruz Biotechnology, Inc, sc-2924) at 1:10000 dilution. Chemiluminescence was detected using PierceTM ECL Western Blotting Substrate and an iBright FL1500 imager (Thermo Fisher Scientific).

#### Transmission Electron Microscopy (TEM)

5μl of VLPs were applied to a formvar/carbon supported 3.05mm copper grid (size 200 mesh) for 10 min before blotting off excess material and negative staining with uranyl acetate for 1 min. Excess stain was removed and grids were air dried for 15 min before imaging on a JEOL-1400 Plus TEM. The approximate diameter of VLPs was measured using the ImageJ line selection tool.

#### Primary B cell culture and stimulation

Mononuclear cells from peripheral blood were isolated as previously described from three COVID-19 naïve donors (N-01, N-02, N-03) (*83*). Briefly, fresh blood samples from healthy individuals were collected in EDTA tubes. Blood was diluted 1:1 with PBS and layered gently on Ficoll-Paque in SepMATE tubes (StemCell Technologies) followed by density gradient centrifugation. Cells were thoroughly washed and were either freshly stained for flow cytometry or were stored in freezing medium (Cryostor CS10 cell cryopreservation media, Cat. C2874, Sigma-Aldrich) at −150°C., frozen PBMC’s were thawed, washed and resuspended in RPMI containing 10% FBS, L-Glutamine, non-essential amino acids, HEPES, and 100U/ml penicillin plus 100ug/ml streptomycin. B cell purification was performed using EasySepTM Human B cell Enrichment Kit (Cat#19054, Stemcell technology) following manufacturer’s protocol. 5×10^4^ cells were plated in 96 u-bottom well plate and stimulated with: (i) VLP 1:10 (10ul VLP resuspension in 100ul total culture media); (ii) VLPs 1:500 (0.2ul in 100ul); (iii) R848 1:250 (Cat#tlrl-r848, Invivogen); (iv) CpGB DNA 1:500 (Cat#HC4039, Cambridge Bioscience); (v) CD40L 10 µg/mL (Cat#6245-CL-050, R&D Systems); (vi) detergent treated VLPs, these were as in (i) but with 1% Triton-x-100 added for 30 mins to dissolve lipid-protein structures before having buffer exchanged to remove detergent by passing over 3kDa MWCO spin column for 6x washes. For unstimulated control, cells were in complete media with no additions. Following stimulation, supernatant was collected, and immunoglobulin profiles were analysed using LegendPlex following manufacturer’s protocol (Cat#740640, Biolegend).

#### Subjects for PBMC donation

Blood was collected for B cell assays under the ethical approval from the Edinburgh Medical School Research Ethics Committee of the University of Edinburgh (21-EMREC-010). Three healthy controls with no known SARS-CoV-2 history, and prior vaccination were recruited as a convenience sample from volunteers at our research institute. After collection of informed consent, demographics, and recent SARS-CoV-2 infection, and SARS-CoV-2 vaccination history was collected. Samples were then pseudonymized at source.

## Data Availability

All data necessary are available in the text or the supplementary materials. Code to reproduce the analyses in the paper can be accessed from the first author's GitHub project repository.

https://github.com/PKKearns/SARS2_Antibody_Profiling_Paper

## Acknowledgments

We are grateful to Professors David Gray and members of the Gilbert Lab and MRC Human Genetics Unit for useful discussions throughout the project. We are grateful to Scott Neilson of the Edinburgh Genome Foundry for assistance with robotic cloning pipelines. We are grateful to the staff of the Scottish National Blood Transfusion Service Microbiology Reference Unit staff for provision of samples from Scottish blood donors. This research was conducted with material produced with the assistance of the Edinburgh Genome Foundry, a synthetic biology research facility specialising in the assembly of large DNA fragments at the University of Edinburgh. We are also grateful to the staff of the IGC, advanced imaging department, for technical assistance particularly Matt Pearson.

Members of the Manchester Coronavirus Immune Response and Clinical Outcomes (CIRCO) consortium: Rohan Ahmed, Miriam Avery, Katharine Birchall, Evelyn Charsley, Alistair Chenery, Christine Chew, Richard Clark, Emma Connolly, Karen Connolly, Simon Dawson, Laura Durrans, Hannah Durrington, Jasmine Egan, Kara Filbey, Claire Fox, Helen Francis, Miriam Franklin, Susannah Glasgow, Nicola Godfrey, Kathryn J. Gray, Seamus Grundy, Jacinta Guerin, Pamela Hackney, Chantelle Hayes, Emma Hardy, Jade Harris, Anu John, Bethany Jolly, Verena Kästele, Gina Kerry, Sylvia Lui, Lijing Lin, Alex G. Mathioudakis, Joanne Mitchell, Clare Moizer, Katrina Moore, Stuart Moss, Syed Murtuza Baker, Rob Oliver, Grace Padden, Christina Parkinson, Michael Phuycharoen, Ananya Saha, Barbora Salcman, Nicholas A. Scott, Seema Sharma, Jane Shaw, Joanne Shaw, Elizabeth Shepley, Lara Smith, Simon Stephan, Ruth Stephens, Gael Tavernier, Rhys Tudge, Louis Wareing, Roanna Warren, Thomas Williams, Lisa Willmore, and Mehwish Younas.

## Author contributions

Conceptualization: PKAK, DJK, JM, NG

Methodology: PKAK, CD, DJK, DC, RK, KL, MM, JM, NG

Computational/bioinformatic analyses: PKAK, MB, OF, LG, JM

Laboratory experiments: PKAK, CD, RC, RS, BP, KL, OF, SB, RK, DJK, NG

Edinburgh cohort: HW, SJ, KT

Oxford cohort: AM, JK, PK, EB, SD, CD, TL, AP

Manchester cohort: MM, TH

Long Covid cohort: AC, LMcW

Synthetic biology pipeline: PKAK, DJK, JG, SN, RF, NG

Statistical analyses: PKAK

Visualization: PKAK, MB

Funding acquisition: PKAK, DJK, TL, PK, EB, SD, KT, JK, AP, JM, NG

Project administration: PKAK, OF, CD, TL, PK, EB, SD, JK, AP, MM, TH, SJ, KT, AC, NG

Supervision: TL, JK, PK, EB, SD, AP, TH, KT, SR, DC, JM, NG

Writing – original draft: PKAK

Writing – review & editing: PKAK, CD, MB, AP, JK, AC, DJK, JAM, NG

All authors reviewed and approved the submitted manuscript.

## Data and materials availability

All data necessary are available in the text or the supplementary materials. Code to reproduce the analyses in the paper can be accessed from the first author’s GitHub project repository (github.com/PKKearns/SARS2_Antibody_Profiling_Paper). Materials generated during the course of this study: aliquots of purified GST-peptide fusion proteins for the prioritized peptides and bacterial/mammalian expression vectors are available on request.

