## Supplement for "Structural epitope profiling identifies antibodies associated with critical COVID-19 and long COVID"

P.K.A. Kearns, et al.

**This PDF file includes:**

Figs. S1 to S20

Tables S1 to S7

Data S1 to S3

| Usage | Product | Vendor | Catalogue number/Product code |
| --- | --- | --- | --- |
| Cloning | DNA E-blocks | IDT | N/A |
|  | BsaI-HF2 | NEB | R3733S |
|  | T4 ligase | NEB | M0202S |
| Protein expression | Ampicillin | Sigma-Aldrich | A9518 |
|  | E. coli NEB 5α competent cells | New England Biolabs | C2987 |
|  | E. coli T7 Express competent cells | New England Biolabs | C2566H |
|  | Benzonase | Merck Millipore | 70664 |
|  | PMSF |  |  |
|  | Expi293F cells | ThermoFisher | A14527 |
|  | Expi293 expression media | ThermoFisher | A1435101 |
| Mammalian expression | ExpiFectamine kit | ThermoFisher | A14525 |
|  | 125ml Erlenmeyer flasks | ThermoFisher | 4115-0125 |
| Protein affinity chromatography | Tween-20 | Sigma-Aldrich | P2287 |
|  | Imidazole | Sigma-Aldrich | 56750 |
|  | Ni-NTA agarose magnetic beads (Pierce) | Thermo Fisher Scientific | 78605 |
|  | Plastics |  |  |
|  | KingFisher Flex robot | Thermo Fisher Scientific | A32681 |
|  | KingFisher deep-well plates (96 and 24 well) | Thermo Fisher Scientific | 95040450  95040470 |
|  | KingFisher Tip Comb (96 and 24) | Thermo Fisher Scientific | 97003500  97002610 |
|  | PD Minitrap G-25 | Cytiva | 28918007 |
|  | PD G-25 MultiTrap Plate | Cytiva | 11793329 |
| Protein Gel | LDS Sample Buffer (4x) | Novex | NP0007 |
|  | Nu-PAGE 4-12% Bis-Tris | Thermo Fisher Scientific | NP0336BOX |
|  | NuPAGE MOPS SDS Buffer (20X) | Thermo Fisher Scientific | NP0001 |
|  | SeeBlue Plus2 (ladder) | Thermo Fisher Scientific | LC5925 |
| ELISA  Antibodies for immunofluorescence and Western blotting | SARS-CoV-2 Spike S1- His Recombinant Protein | Sino Biological | 40591-V08H |
|  | 96 Well ELISA  Microplate, PS,  MICROLON®,  F-Bottom | Greiner bio-one | 655001 |
|  | Rabbit Anti-Human  IgG, Polyclonal, HRP | Dako, Agilent Technologies | P0214 |
|  | Rabbit Anti-Human  IgA, Polyclonal, HRP | Dako, Agilent Technologies | P0216 |
|  | Rabbit Anti-Human  IgM, Polyclonal HRP | Dako, Agilent Technologies | P0215 |
|  | Multichem IDCOVID19Neg | Technopath Clinical  Diagnostics | CVN200N |
|  | Multichem IDCOVID19 G | Technopath Clinical  Diagnostics | CVG200P |
|  | Sheep Anti-SARS-CoV-2 M protein, Polyclonal | MRC PPU Reagents and Services | DA107 |
|  | Sheep Anti-SARS-CoV-2 E protein, Polyclonal | MRC PPU Reagents and Services | DA108 |
|  | Sheep Anti-SARS-CoV-2 N protein, Polyclonal | MRC PPU Reagents and Services | DA114 |
|  | Rabbit Anti-Sheep, Polyclonal HRP | Santa Cruz Biotechnology, Inc. | sc-2924 |
|  | Donkey Anti-Goat IgG, Alexafluor Plus 594 | Thermo Fisher Scientific | A32758 |

R packages and version numbers used in analyses

Base packages:

stats	graphics grDevices utils	datasets methods	base

Other packages:

Peptides_2.4.4	ggcorrplot_0.1.3	GGally_2.1.2	ggfortify_0.4.14	jtools_2.2.0	ggpmisc_0.4.6	ggpp_0.4.4	mgcv_1.8-40	nlme_3.1-157	broom.mixed_0.2.9.4 sjstats_0.18.1	effects_4.2-1	sjmisc_2.8.9	sjPlot_2.8.10	MuMIn_1.46.0	lmerTest_3.1-3	officer_0.4.2	gridExtra_2.3	patchwork_1.1.1	cowplot_1.1.1	bayestestR_0.11.5	lubridate_1.8.0	performance_0.9.0	equatiomatic_0.3.1 yardstick_0.0.9	workflowsets_0.2.1 workflows_0.2.6	tune_0.2.0	rsample_0.1.1	recipes_0.2.0	parsnip_0.2.1	modeldata_0.1.1	infer_1.0.0	dials_0.1.1	broom_0.8.0	tidymodels_0.2.0	flextable_0.7.0	gtsummary_1.6.0	styler_1.7.0	lme4_1.1-29	Matrix_1.4-1	writexl_1.4.0	umap_0.2.7.0	cutpointr_1.1.2	viridis_0.6.2	viridisLite_0.4.0	growthcurver_0.3.1 ggpubr_0.4.0	ggrepel_0.9.1	gtools_3.9.2	plater_1.0.4	magrittr_2.0.3	drc_3.0-1	MASS_7.3-56	scales_1.2.0	naniar_0.6.1	reshape2_1.4.4	readxl_1.4.0	car_3.0-12	carData_3.0-5	forcats_0.5.1	stringr_1.4.0	dplyr_1.0.8	purrr_0.3.4	readr_2.1.2	tidyr_1.2.0	tibble_3.1.6	ggplot2_3.3.5	tidyverse_1.3.1	pacman_0.5.1

Loaded via a namespace (not attached):

estimability_1.3	SparseM_1.81	R.methodsS3_1.8.1	coda_0.19-4	visdat_0.5.3	knitr_1.39	multcomp_1.4-19	R.utils_2.11.0	data.table_1.14.2	rpart_4.1.16	hardhat_0.2.0	doParallel_1.0.17	generics_0.1.2	GPfit_1.0-8	BiocGenerics_0.42.0	TH.data_1.1-1	future_1.25.0	tzdb_0.3.0	xml2_1.3.3	httpuv_1.6.5	assertthat_0.2.1	gower_1.0.0	xfun_0.30	hms_1.1.1

evaluate_0.15	promises_1.2.0.1	fansi_1.0.3	dbplyr_2.1.1	DBI_1.1.2	reshape_0.8.9	stats4_4.2.0	ellipsis_0.3.2	RSpectra_0.16-1	backports_1.4.1	insight_0.17.0	survey_4.1-1	vctrs_0.4.1	quantreg_5.88	here_1.0.1	sjlabelled_1.2.0	abind_1.4-5	withr_2.5.0	emmeans_1.7.3	cluster_2.1.3	crayon_1.5.1	pkgconfig_2.0.3	nnet_7.3-17	rlang_1.0.2	globals_0.14.0	lifecycle_1.0.1	MatrixModels_0.5-0	sandwich_3.0-1	modelr_0.1.8	cellranger_1.1.0	rprojroot_2.0.3	matrixStats_0.62.0	datawizard_0.4.0	boot_1.3-28	zoo_1.8-10	reprex_2.0.1	base64enc_0.1-3	GlobalOptions_0.1.2	png_0.1-7	rjson_0.2.21	parameters_0.17.0	R.oo_1.24.0	pROC_1.18.0	pander_0.6.5	shape_1.4.6	parallelly_1.31.1	R.cache_0.15.0	rstatix_0.7.0	ggeffects_1.1.2	S4Vectors_0.34.0	ggsignif_0.6.3	plyr_1.8.7	compiler_4.2.0	RColorBrewer_1.1-3	plotrix_3.8-2	clue_0.3-60	cli_3.3.0	DiceDesign_1.9	listenv_0.8.0	tidyselect_1.1.2	stringi_1.7.6	mitools_2.4	askpass_1.1	grid_4.2.0	tools_4.2.0	future.apply_1.9.0	parallel_4.2.0	circlize_0.4.14	rstudioapi_0.13	uuid_1.1-0	foreach_1.5.2	prodlim_2019.11.13	digest_0.6.29	BiocManager_1.30.17	shiny_1.7.1	ggtext_0.1.1	lava_1.6.10	Rcpp_1.0.8.3	gridtext_0.1.4	later_1.3.0	httr_1.4.2	gdtools_0.2.4	ComplexHeatmap_2.12.0 effectsize_0.6.0.1	colorspace_2.0-3	rvest_1.0.2	fs_1.5.2	reticulate_1.24	IRanges_2.30.0	splines_4.2.0	systemfonts_1.0.4	xtable_1.8-4	jsonlite_1.8.0	nloptr_2.0.0	timeDate_3043.102	ipred_0.9-12	gt_0.5.0	R6_2.5.1	lhs_1.1.5	pillar_1.7.0	htmltools_0.5.2	mime_0.12	glue_1.6.2	fastmap_1.1.0	minqa_1.2.4	class_7.3-20	codetools_0.2-18	mvtnorm_1.1-3	furrr_0.2.3	utf8_1.2.2	lattice_0.20-45	numDeriv_2016.8-1.1 zip_2.2.0	openssl_2.0.0	survival_3.3-1	rmarkdown_2.14	munsell_0.5.0	GetoptLong_1.0.5	iterators_1.0.14	broom.helpers_1.7.0 haven_2.5.0	gtable_0.3.0

Supplementary Figures

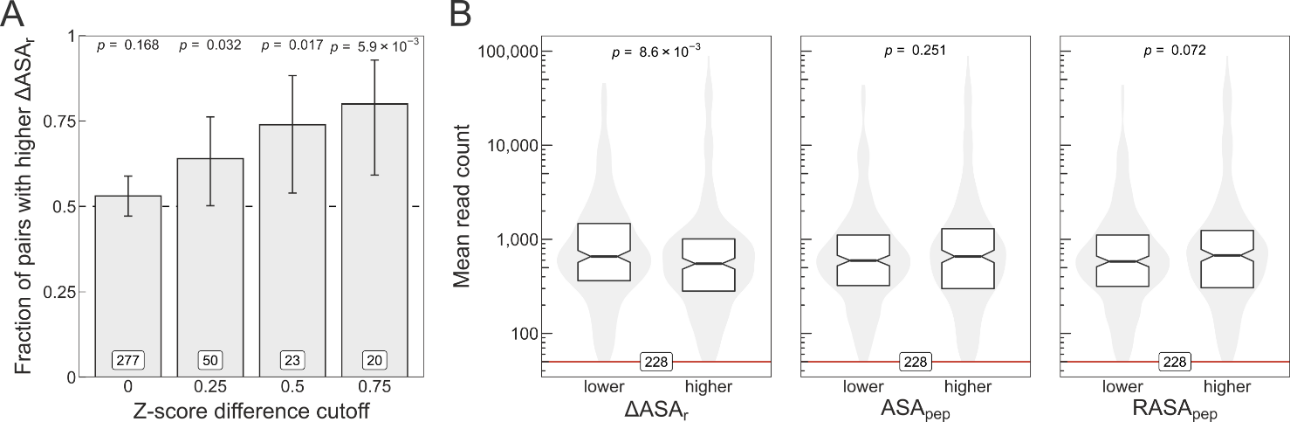

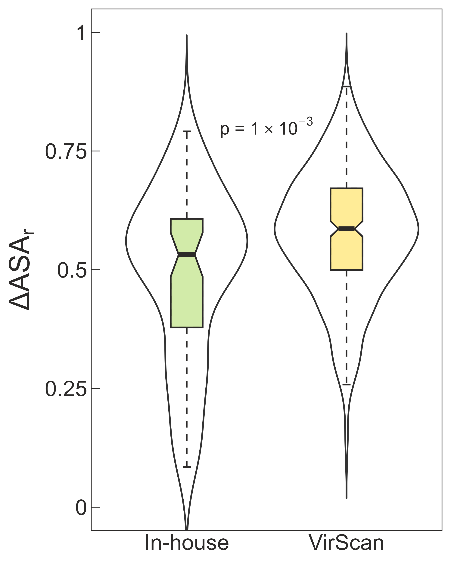

Fig. S1. Structural information captured by ΔASAr could be improve library design in high-throughput platforms. (A) Analysis of overlapping VirScan peptides. Bars represent the fraction of peptide pairs that exhibit a higher ΔASA_r_ and have a higher immunogenicity score. The X-axis represents Z-score difference cutoffs of immunogenicity scores, *i.e.* the higher the cutoff the more the peptides were found enriched for binding in the VirScan data. Error bars are 95% Jeffrey’s confidence intervals, labels on bars indicate the number of pairs at the given Z-score cutoff, and the *p*-values were calculated with a one-sided binomial test. (B) Boxplot/violin-plot analysis of adjacent ReScan peptides. Read counts from phage display were averaged across patients for each peptide and adjacent peptides were paired according to “lower” and “higher” values of ΔASA_r_ as well as the solvent-accessible surface area of the peptide (ASA_pep_) and the relative solvent-accessible surface area of the peptide (RASA_pep_). The ΔASA_r_ parameter, but not ASA_pep_ or RASA_pep_, was found to be significantly predictive of peptide immunogenicity. The *p*-values were determined using the two-sided Wilcoxon signed-rank test. Red line indicates the minimum read count considered for a peptide (=50), and labels at the bottom show the number of peptide pairs analysed. (C) Data as in Fig 1E for the virus Spike peptides we show that the distribution of ΔASAr from our chosen peptides is lower than the VirScan peptides. As per figure S3, we show that using VirScan or ReScan data only, where overlapping peptides are compared the peptide with the lower ΔASAr is more likely to be immunogenic. This implies that there is substantial scope for high-throughput methods optimizing their peptide selection using the ΔASAr.

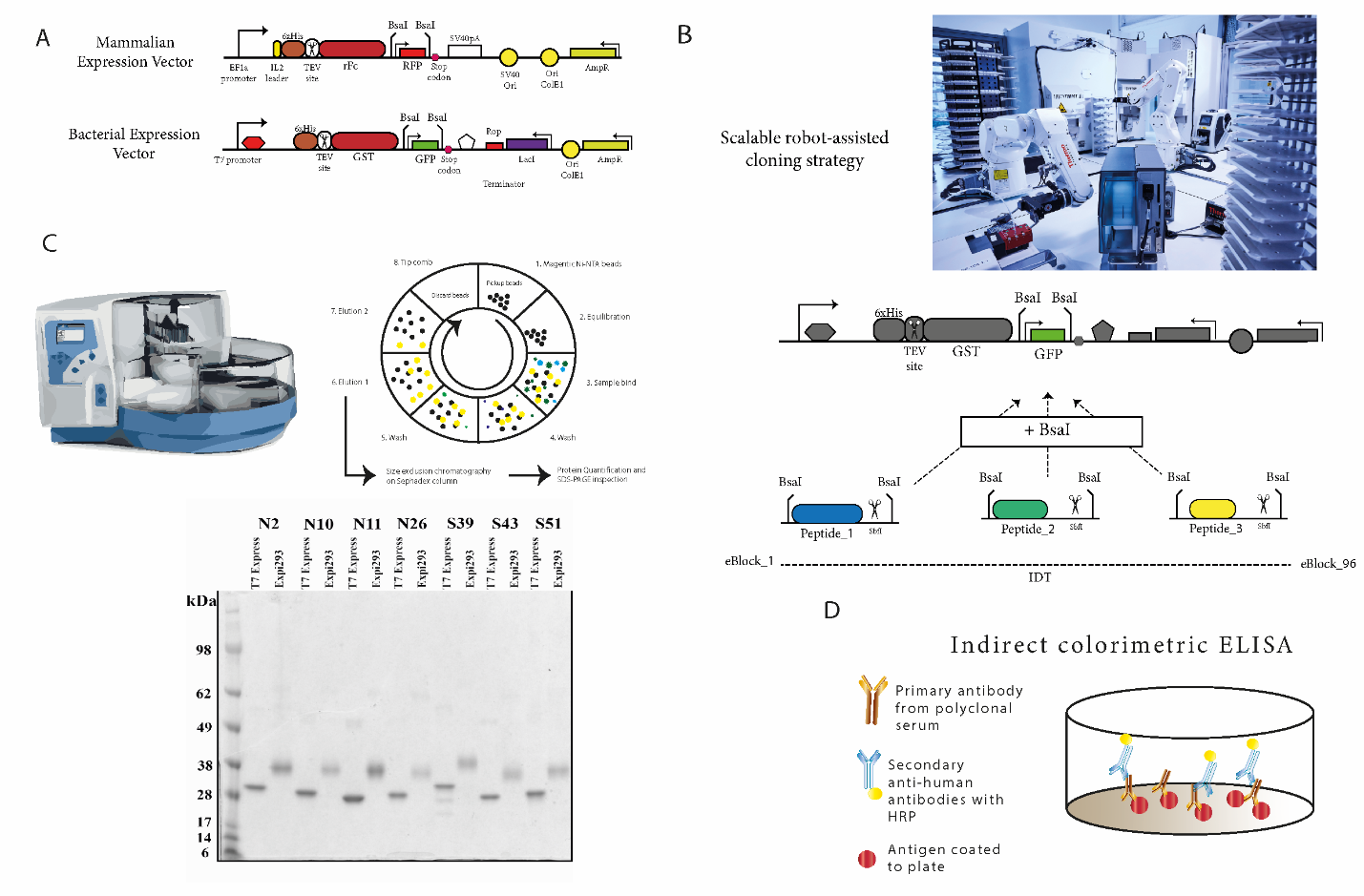

Fig. S2. Design and optimization of a synthetic biology pipeline for scalable cloning and protein expression. (A) Schematic of novel mammalian and bacterial expression vectors. EF1a promoter, TEV – Tobacco Etch Virus protease cleavage site, RFP – red fluorescent peptide, SV40pA – SV40 poly-A tail, SV40 origin of replication, COlE1 origin of replication, ampicillin resistance cassette, 6xHis – poly-histidine affinity tag, GST – glutathione S-transferase, GFP – green fluorescent protein, LacI – Lac repressor protein, ROP – repressor of ColE1 primer. (B) golden gate cloning using commercially synthesized DNA blocks and directional Type IIS restriction enzyme sites (e.g. BsaI). (C) Robot assisted high throughput cloning, colony picking, diagnostic restriction enzyme digest, Sanger sequencing, bacterial transformation, lysis, and Ni-NTA affinity chromatography protein purification, buffer exchange and quantification pipeline was established and optimized. Yielding reliable clean fusion peptides (representative examples on SDS PAGE 4-12% Bis-Tris gel run in MOPS SDS buffer. “T7 Express” labelled are *E. coli* expressed and “Expi293” are mammalian expressed fusion proteins. (D) Schematic illustrating principle of fusion protein indirect colorimetric ELISAs in a single well of a 96 well plate.

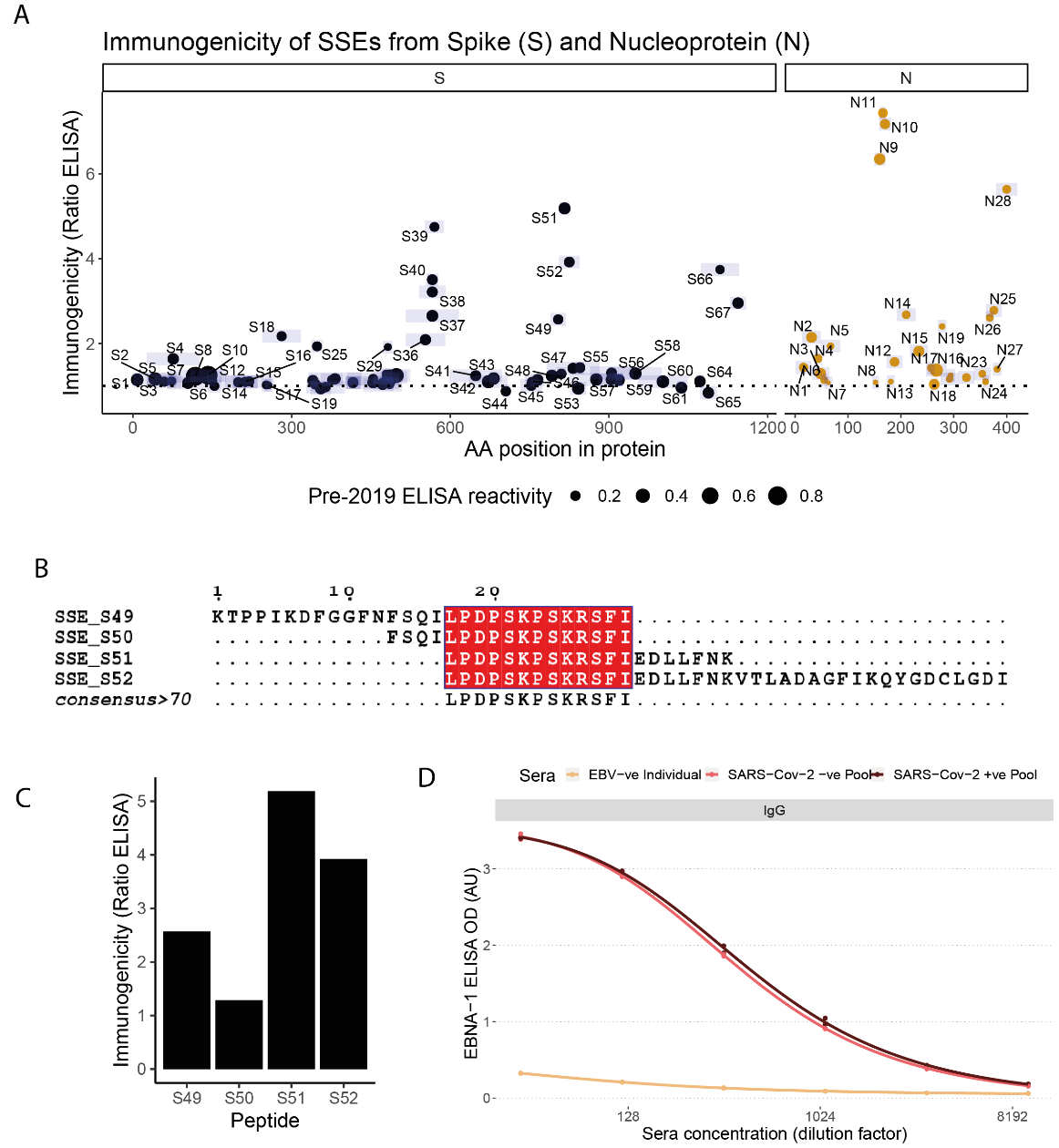

Fig. S3. Immunogenicity of N and S peptides and position on protein illustrating overlapping peptides. (A) Immunogenicity of structurally stable epitopes (SSEs) from the spike (S) and nucleoprotein (N) of SARS-CoV-2 as expressed in the bacterial system. Y-axis is the ratio of mean of at least triplicate values comparing pooled SARS-CoV-2 convalescent sera to pooled pre-2019 sera (unexposed). Higher values on the y-axis suggest that peptides are more immunogenic. Each point is a peptide, x-axis position marks the central amino acid of the peptide, point size represents the reactivity in the unexposed sera. Shaded boxes give the size of the peptide. Immunodominant SSEs identified in three regions of the spike (S39, S51, S67) and multiple domains of the nucleoprotein (N2, N11, N12, N14, N19, N26, and N28). (B) Multiple sequence protein alignment of overlapping predicted SSEs. (C) Ratio of immunogenicity by peptide is non-obvious based on sequence alone. (D) The pre-2019 sera and post-COVID sera show almost identical titres to an immunodominant Epstein Barr Virus latent antigen (EBNA-1), demonstrating that differences in immunogenicity ratio for SARS-CoV-2 peptides are not due to degradation of the pre-2019 sera.

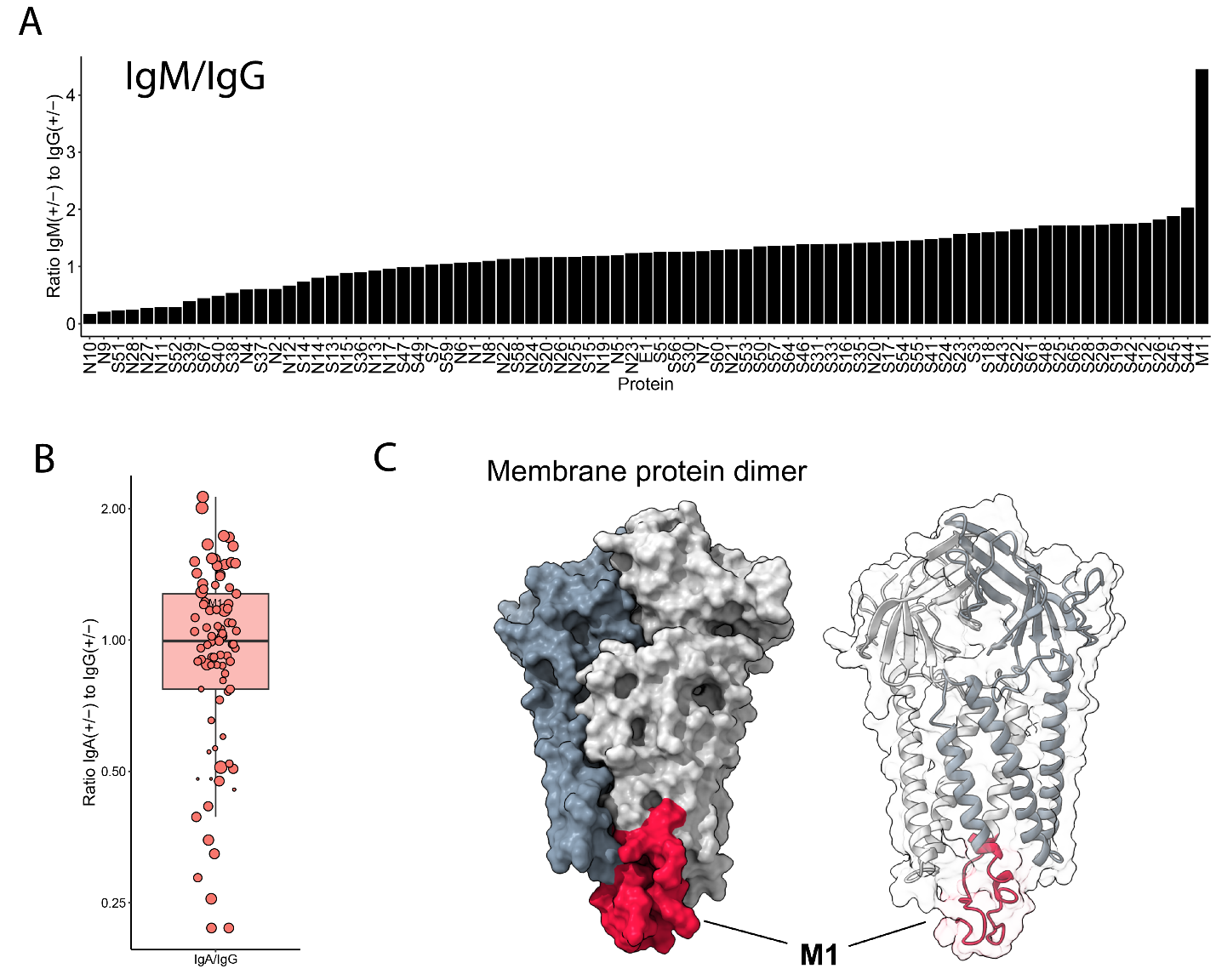

Fig. S4. IgM/IgG for structural peptides. (A) IgA immunogenicity in bacterially expressed proteins as in Fig 2A for IgG and IgM. (B) Ratio of IgA to IgG reactivity demonstrating some epitopes are enriched in IgA vs IgG response and vice versa. (C) Surface and ribbon diagram showing the organization of M proteins within a homodimer and the position of the M1 epitope.

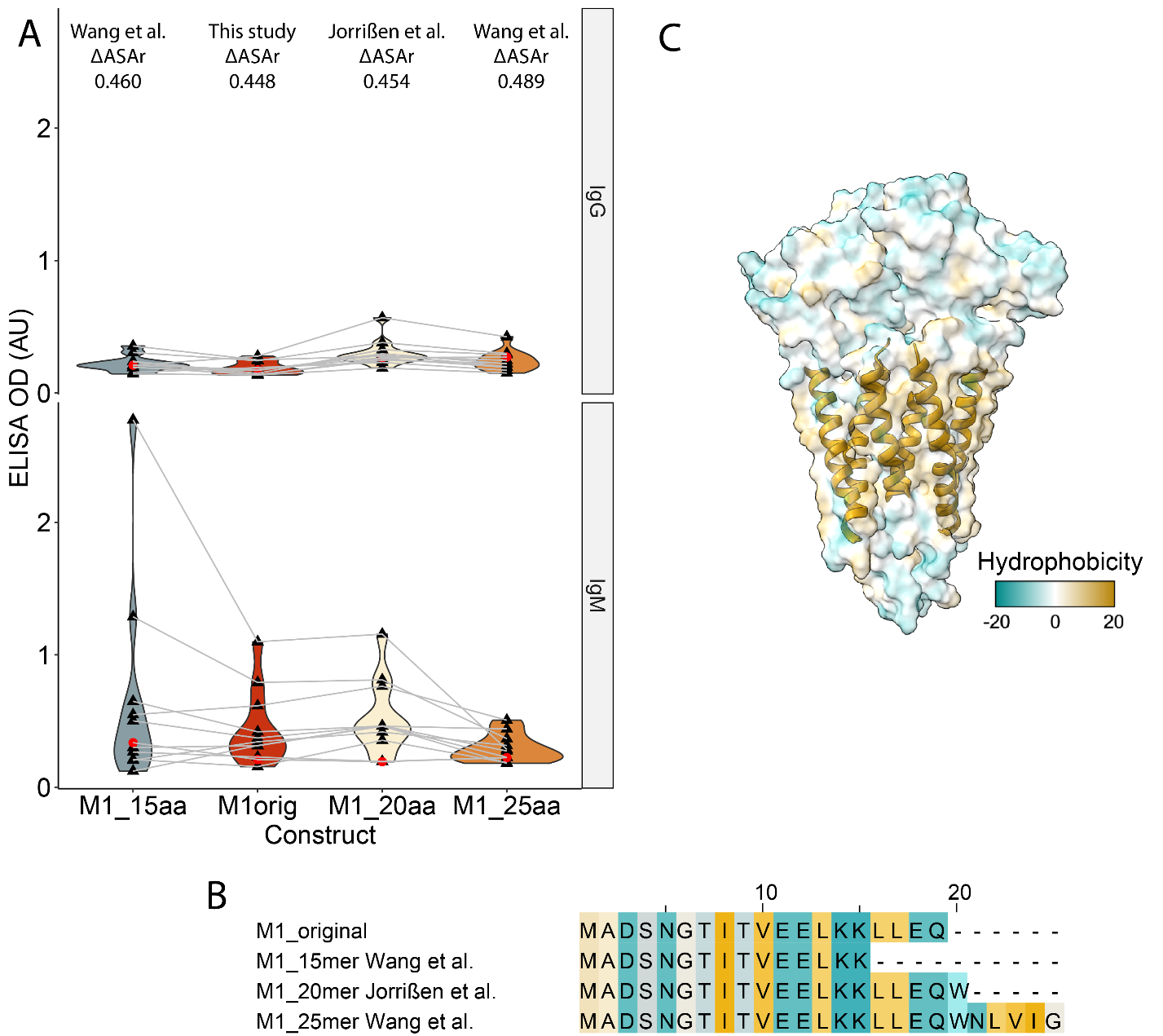

Fig. S5. Design of M1 peptide affects immunogenicity. (A) M1 constructs of varying lengths from peptide micro-array studies show that extending the peptide to include hydrophobic residues (as per 25-mer M1 construct from Wang et al.) reduces immunogenicity. Data points are means of replicates from individuals recovered from SARS-CoV-2 infection (SNBTS donors - black) or from a pool of SARS-CoV-2 naïve individuals from pre-2019 (red). Using peptides that overlap our M1 peptide from two peptide microarray studies (Wang et al. and Jorrigen et al.) we show that, consistent with their data, the longest peptide is the least reactive and may have higher background non-specific reactivity (presumed from IgG reactivity to the negative sera). Our method predicts that the inclusion of the extra six amino acids, would diminish stability of the peptide, reflected by the increased ΔASAr. The 20aa construct performs similarly to our 19aa original peptide (M1orig). The 15aa construct is variable, with some individuals having higher, and some individuals having lower apparent reactivity than the M1orig, and M1_20aa peptides. (B) Structure of the M dimer with surface coloured by Kyte-Doolittle hydrophobicity of amino acids. The ribbon for the transmembrane domain is shown. (C) Alignment of peptides in *A* coloured as *B*.

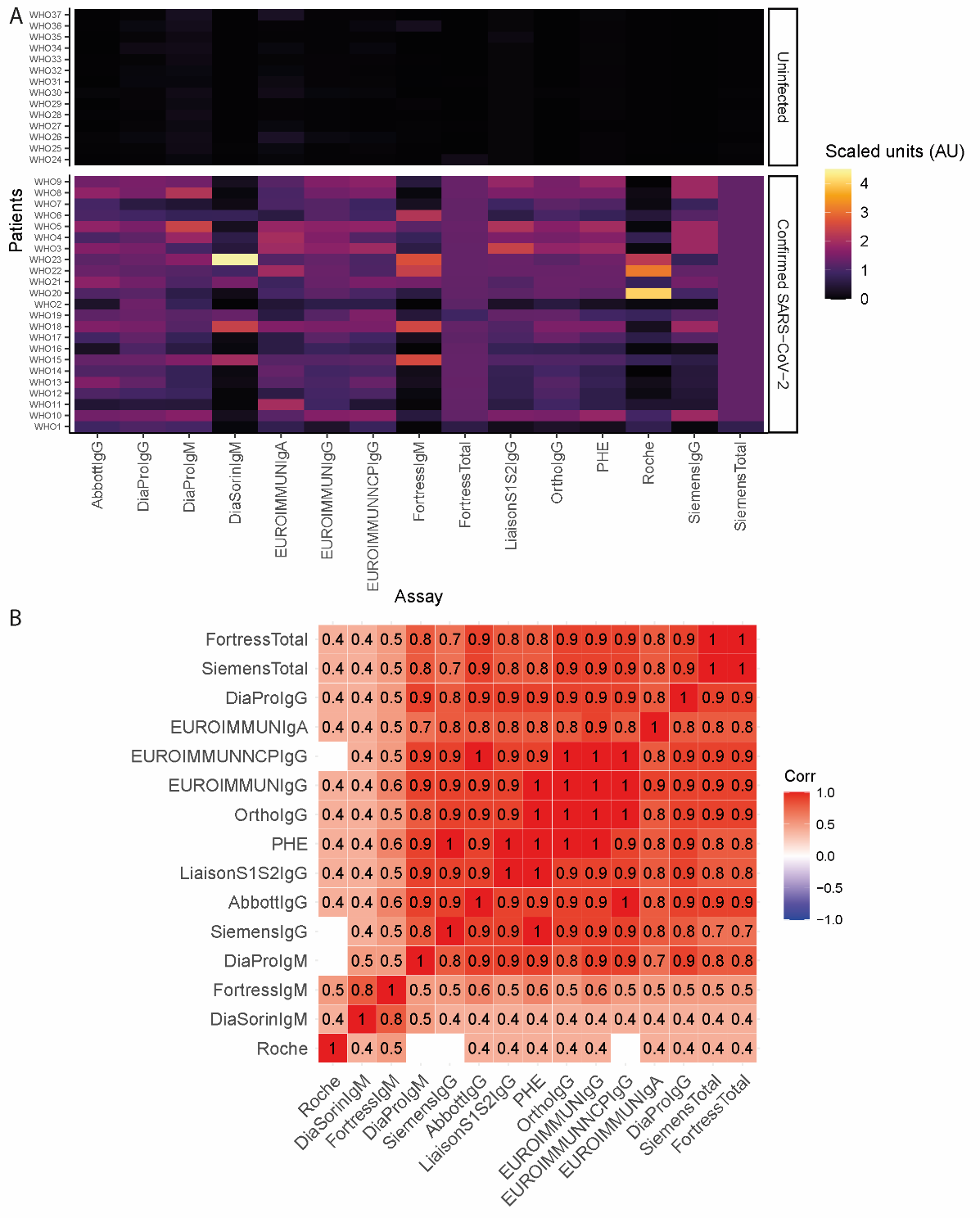

Fig. S6. Commercial assay data for the NIBSC reference sera demonstrating aggregate responses against whole proteins do not show much heterogeneity between subjects in contrast to single epitopes. (A) Top panel uninfected subjects and bottom panel recovered individuals. (B) Correlation matrix of data in A demonstrating that most commercial assays are highly correlated despite the heterogeneity in the individual response at the epitope level due to aggregating these effects.

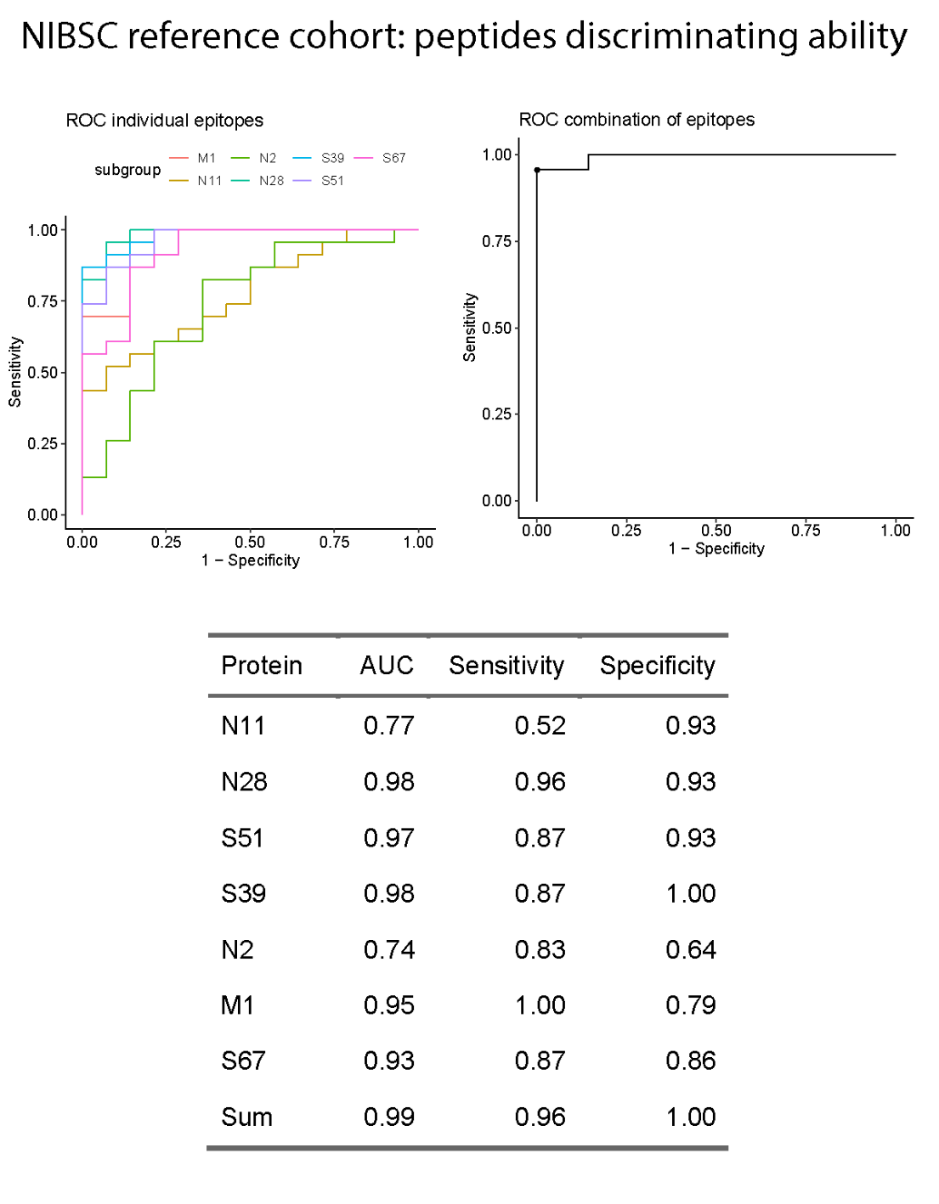

Fig. S7. Receiver operator curves (ROC) for 7 predictive epitopes and the combination in response across in discriminating between infected and naïve individuals.

ROC curves of individual peptides and the combination of signal between them. Table demonstrating the area under the curve (AUC) and sensitivity and specificity for individual and the combination (sum) of responses demonstrating good discriminating ability.

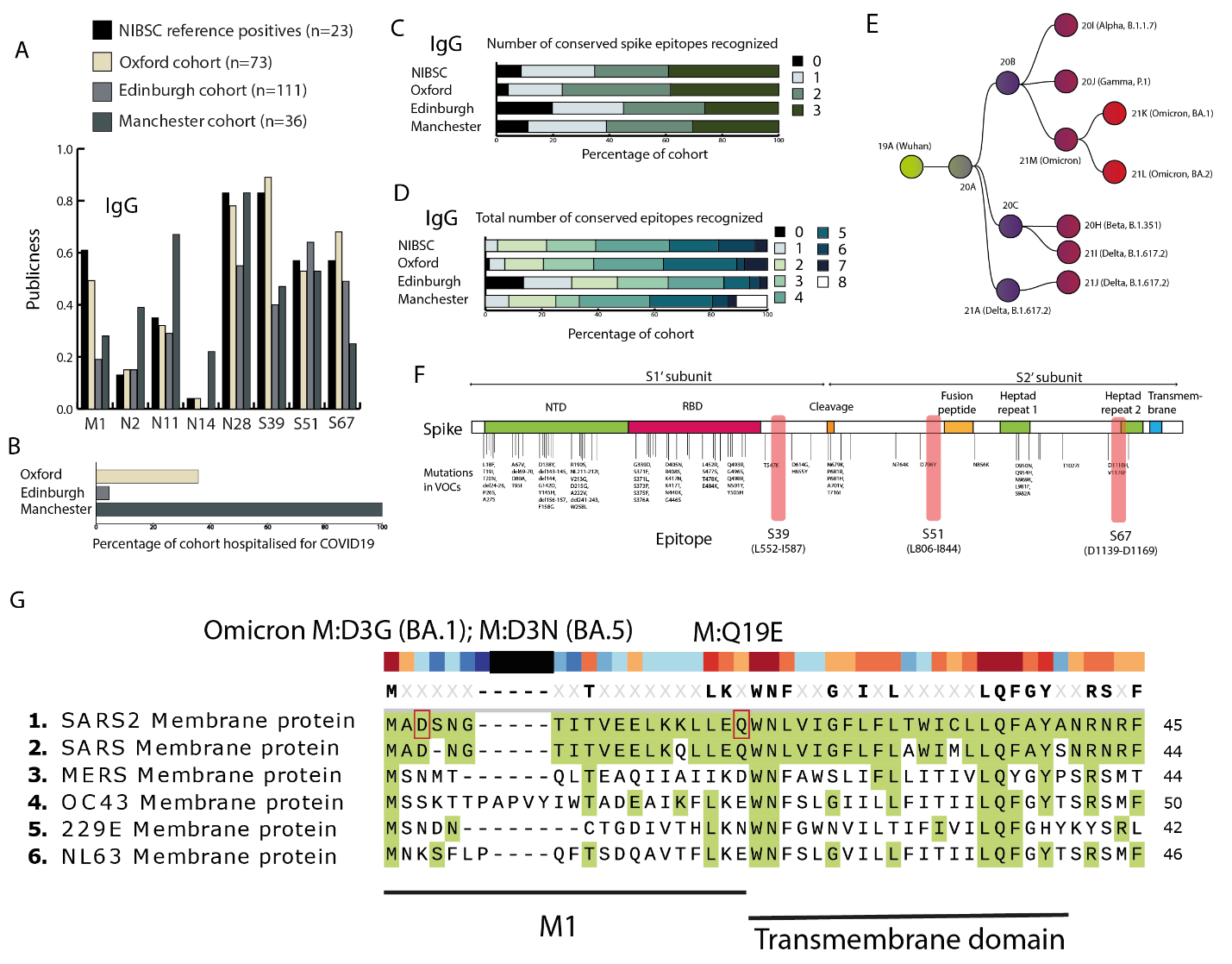

**Fig. S8.** **Antibody responses to the identified epitopes are heterogenous between individuals and epitopes are conserved through virus evolution to date.** (**A**) Comparison of the proportion or “publicness” of individuals in each cohort with positive antibody responses. A positive antibody response was defined as a response >2 SDs from the mean of the NIBSC unexposed cohort. All cohorts were individuals from the pre-vaccination era infected early in 2020. Demonstrating that some epitopes are more public than others but that proportions of individuals reacting between cohorts is similar. (B) Proportion of each cohort hospitalised for COVID-19. (C) Number of spike epitopes recognised by individuals in the cohorts (IgG). (D) Number of eight selected epitopes (as in A) recognized by individuals (IgG). (E) Phylogeny of the SARS-CoV-2 variants of concern. (F) Spike gene map with mutations from variants in E marked on the map with position of epitopes (S39, S51, and S67) annotated. NTD – N-terminus domain and RBD – Receptor binding domain. (G) Multiple sequence alignment (Muscle) of the N-terminus of the membrane protein with other human infecting coronaviruses. Location of the D3 and M19 mutations in Omicron subvariants annotated (red boxes).

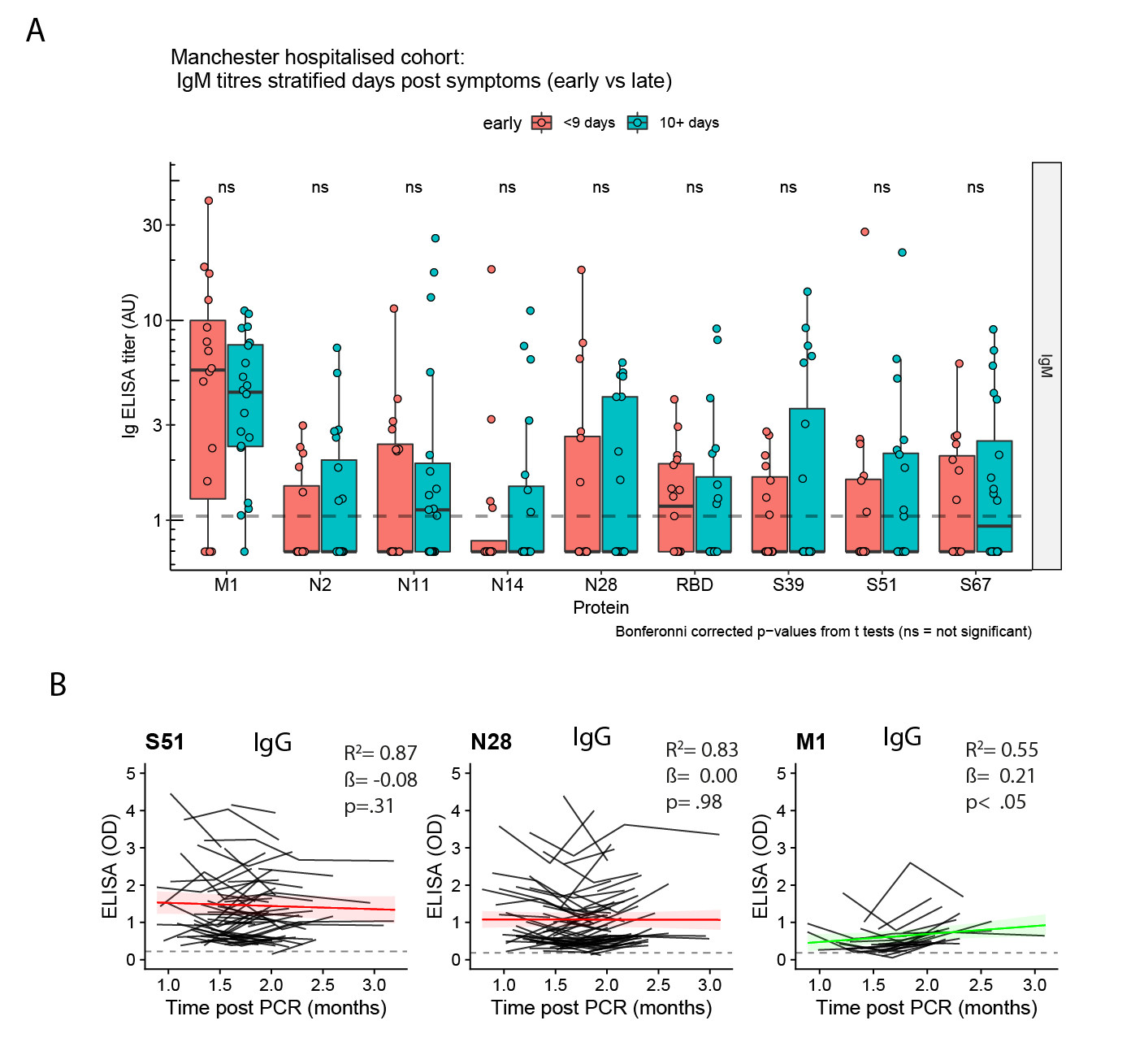

Fig. S9. IgM titres in Manchester cohort of hospitalised patients early in course of disease. (A) Most individuals have detectable IgM against the M1 peptide early (within a week) of symptom onset. Dichotomising this cohort into early and later recruitment post-symptom onset shows that most individuals have detectable IgM titres early in infection, and this is the only epitope where the majority of individuals have detectable titers (B) Longitudinal samples IgG for Edinburgh cohort as summarized in Fig 3E. Shown are individuals who have at least one positive result across study visits. Mixed effects random intercept models are fitted to determine the trends in trajectory.

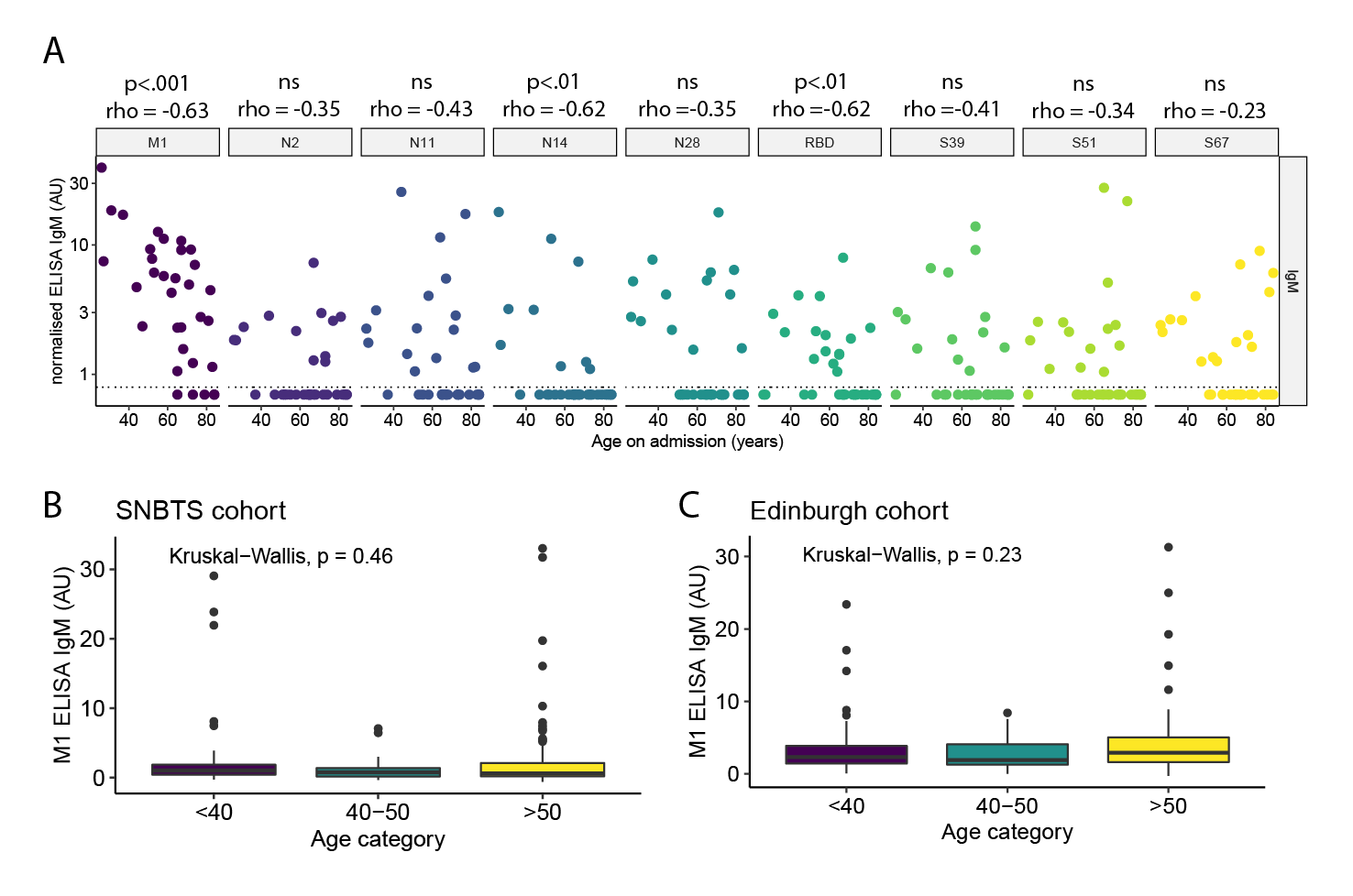

Fig. S11. Effect of days post symptom onset and age at admission association with M1 IgM. (A) In the Manchester (early post-infection, hospitalised) cohort young persons have significantly higher IgM titres in peripheral blood consistent with a slower secretory response in older persons. This may be a general phenomenon of lower virus-specific IgM associated with age as a similar significant correlation exists for the whole receptor binding domain, which is an aggregate of many epitope sites. Spearman’s Rho correlation coefficient with bonferonni-corrected p-value (ns = not significant (corrected p-value ≥ 0.05). (B) The association between age and M1 IgM is not observed in (B) the SNBTS cohort (n=220, 2-6 months post infection) or (C) in the convalescent phase of infection in the first sample of longitudinal Edinburgh cohort (n=111, days post PCR+ 34-46).

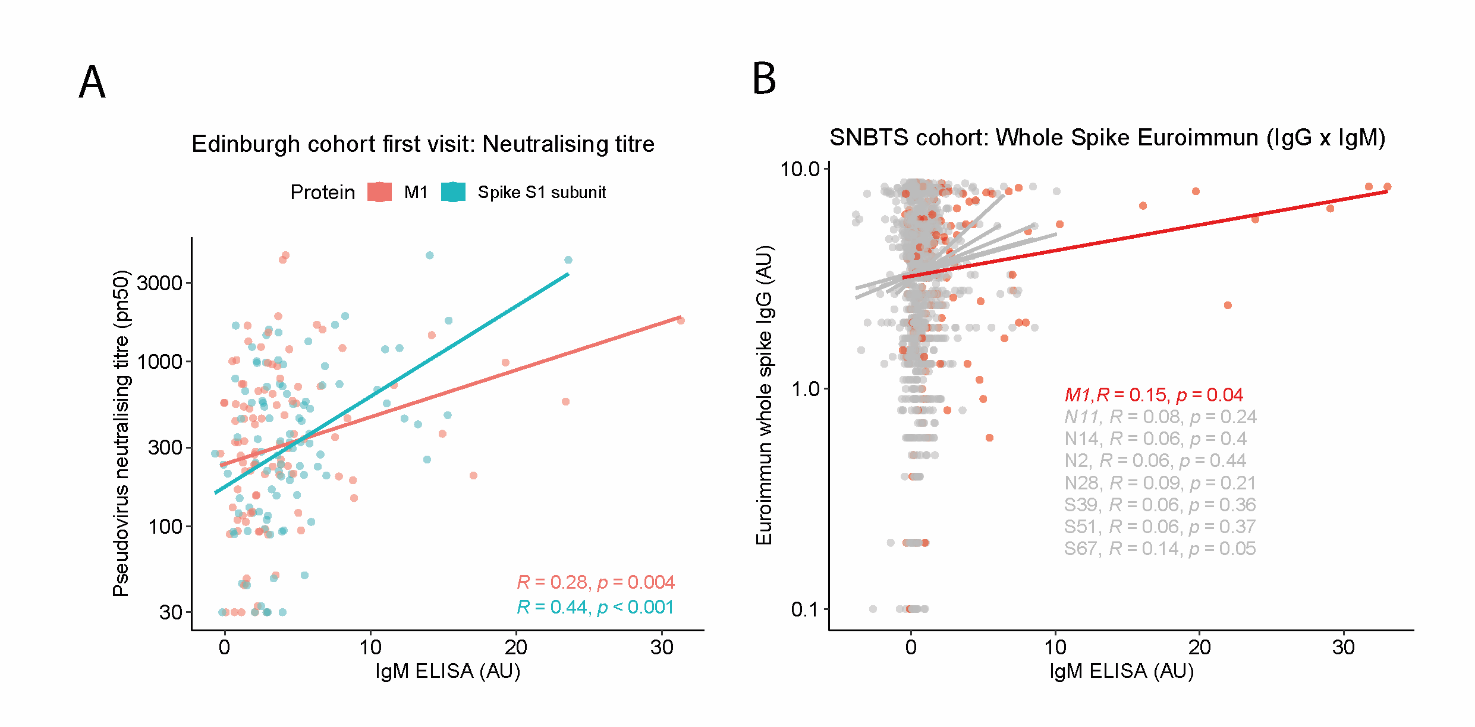

Fig. S12. M1 IgM predicts anti-spike IgG titres and pseudovirus neutralization titres almost as well as IgM to whole S1’ subunit of the spike protein (the site of most neutralizing antibodies). (A) Linear regression of pseudovirus neutralization titre (pn50) against M1 IgM and Spike S1’ subunit. (B) Replication of the predictive ability of IgM M1 in covalescence predicting whole spike IgG in Scottish blood donors (SNBTS).

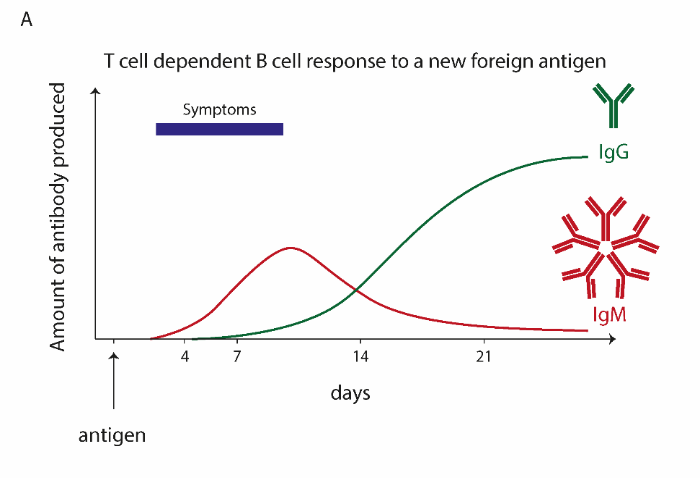

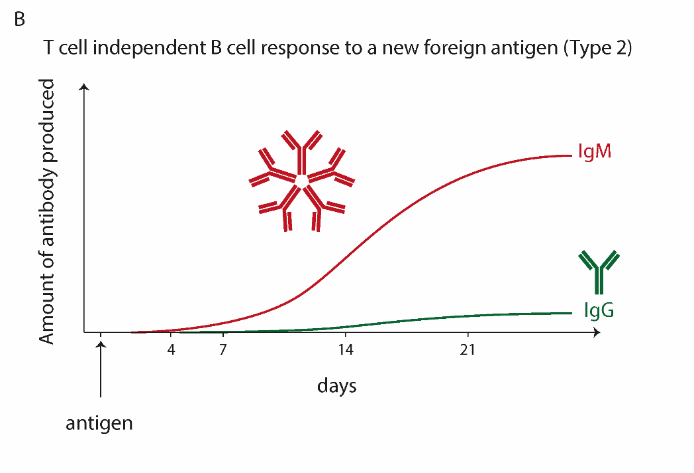

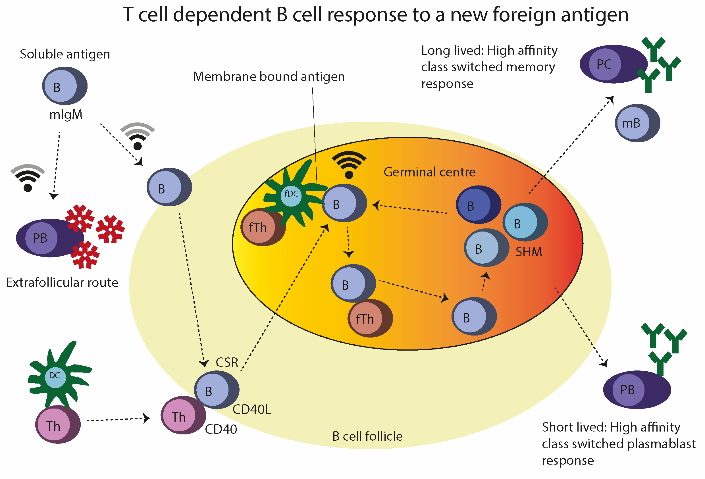

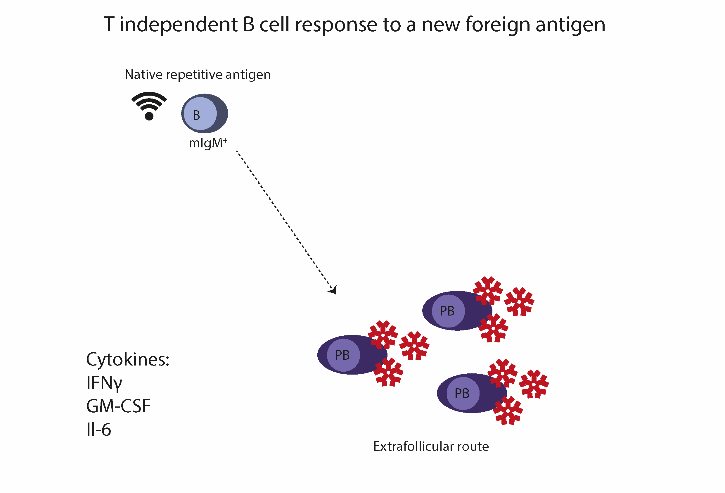

Fig. S13. Typical patterns of antibody production in T dependent and T independent antigen specific immune responses. (A) T cell dependent antibody responses to a novel antigen are characterised by early expansion of IgM^+^ B cells via the extrafollicular plasmablast route and early (starting day 3) class switch recombination to IgG (and other isotypes). (B) In a T independent antibody response, signaling strength of the B cell receptor is strong enough that the extrafollicular plasmablast route is dominant and soluble IgM dominates IgG throughout the response with little class switching and no affinity maturation (which requires somatic hypermutation and rounds of selection in germinal centres). (C) Within secondary lymphoid tissue, B cells that have encountered their cognate antigen are either activated by the extrafollicular route and differentiate into short-lived IgM-secreting plasmablasts, or enter B cell follicles where they receive T cell help at the B cell: T cell border. Here they are stimulated by CD4^+^ T cells via CD40/CD40L and other receptors which stimulates isotype class switch recombination (CSR). B cells then migrate into germinal centres, where they are further stimulated by encountering clustered cognate antigen presented on the membranes of follicular dendritic cells (fDC) and follicular T helper cells (fTh). They undergo cycles of somatic hypermutation and selection based on the affinity of their mutated variable regions of their B cell receptors for antigen and finally they differentiate into long lived plasma cells (PC) or short lived high affinity class switched plasmablasts (PB) or memory B cells (mBC) via a complex interaction with regulatory lymphocytes which are thought to screen out potentially autoreactive clones. (D) In contrast, in response to a T independent antigen the repetitive structure of the antigen in its native form essentially yields a similar signal strength as would be achieved by a presentation on follicular dendritic cells. Local clusters of B cell receptors are cross-linked leading to extensive clonal expansion and plasmablast differentiation with limited class switch recombination, no affinity maturation, and limited or no regulatory checks to prevent autoreactivity.

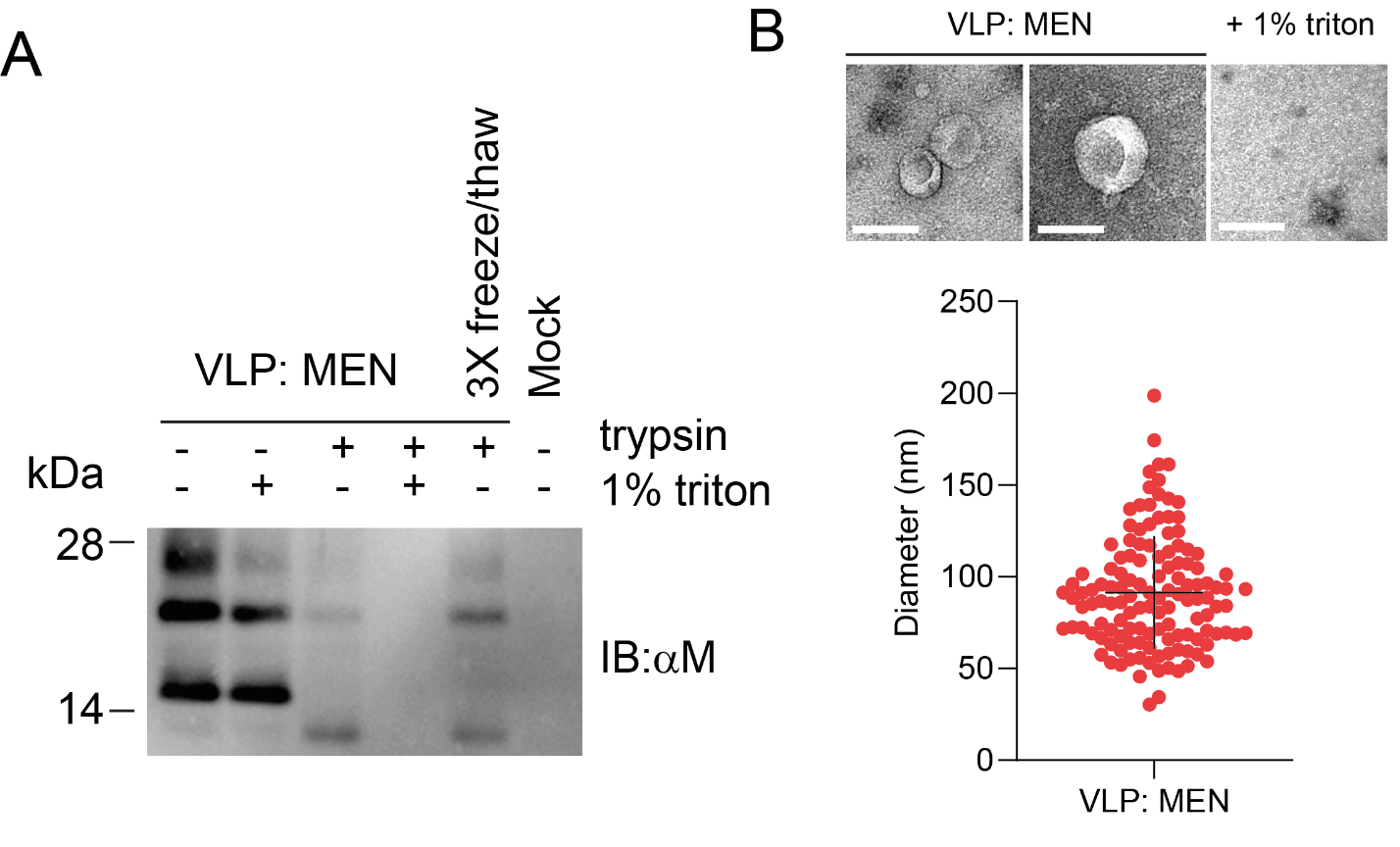

Fig. S14. Characterisation of VLPs. (A) Protease protection assay: Western Blot for M protein in purified VLPs showing protease resistance in the absence of detergent. IB:αM immunoblot with polyclonal sheep anti-M protein antibody. (B) Transmission Electron-Microscopy (TEM) validation of the production and expected size of spikeless VLPs.

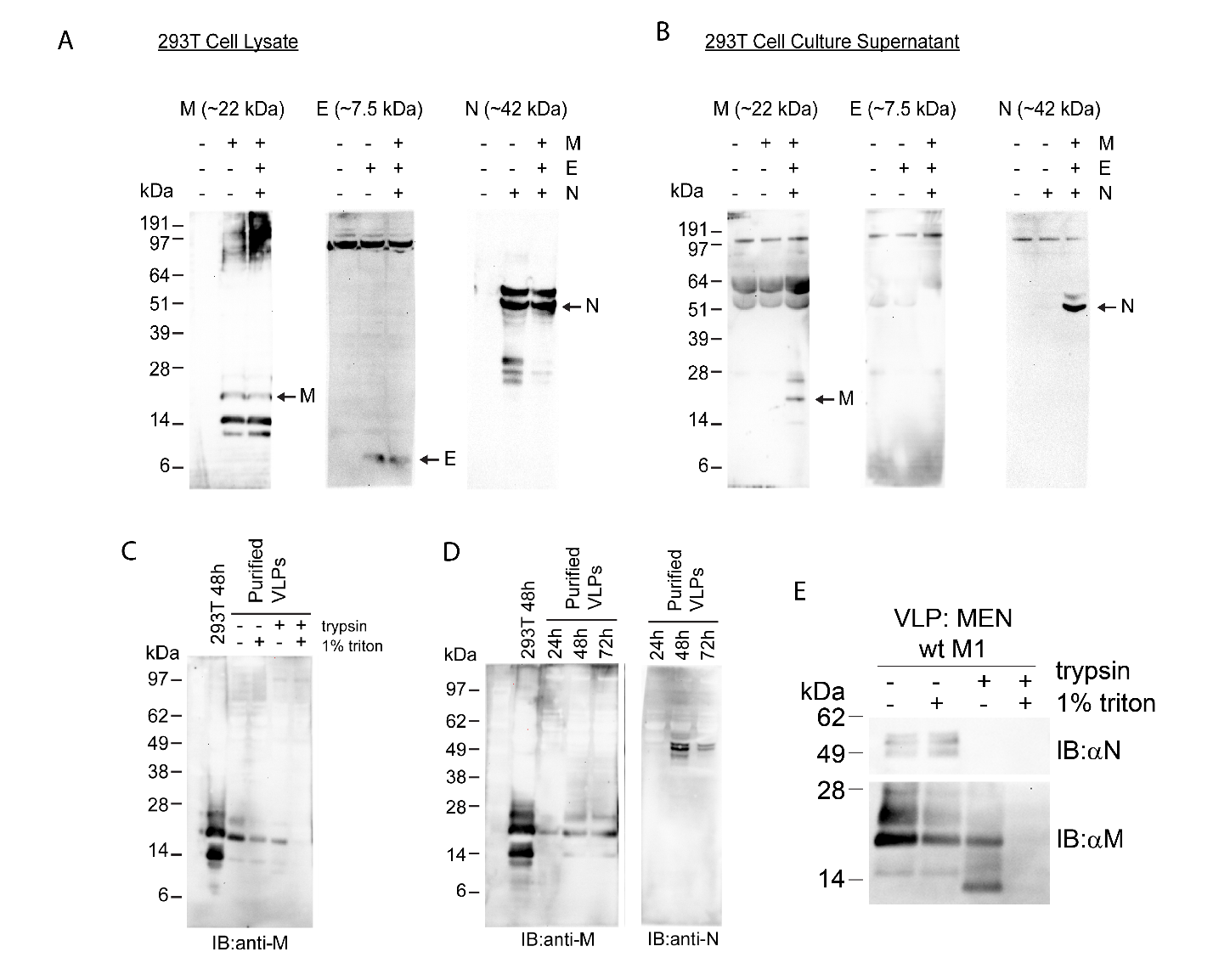
Fig. S15. Characterisation of SARS-CoV-2 spikeless VLPS. (A) Western blots from 293T cell lysate. (B) Western blots from culture supernatant from experiment in A. Background bands can be seen in M blot from low level cross-reactivity with very abundant BSA which is carried through purification on sucrose cushions as some BSA forms large aggregates, this prompted us to switch to produce VLPs in serum free conditions using Optimem which unexpectedly improved yield in addition to reducing background protein contamination. (C) First column is untreated 293T cell lysate harvested at 48hours, subsequent columns are VLPs collected in supernatant purified on a 20% sucrose cushion by ultracentrifugation incubated +/- 1% triton x-100 and/or trypsin in PBS, demonstrating protection of the majority of the M protein in the absence of detergent consistent with presence in intact VLPs. (D) Yield from culture supernatant at 24, 48, and 72hours; blot for M and N. (E) Notably, consistent with most other reports, we found that most secreted N was exposed to trypsin digest and apparently was either not packaged in VLPs, or had accumulated in the supernatant from the degradation of VLPs.

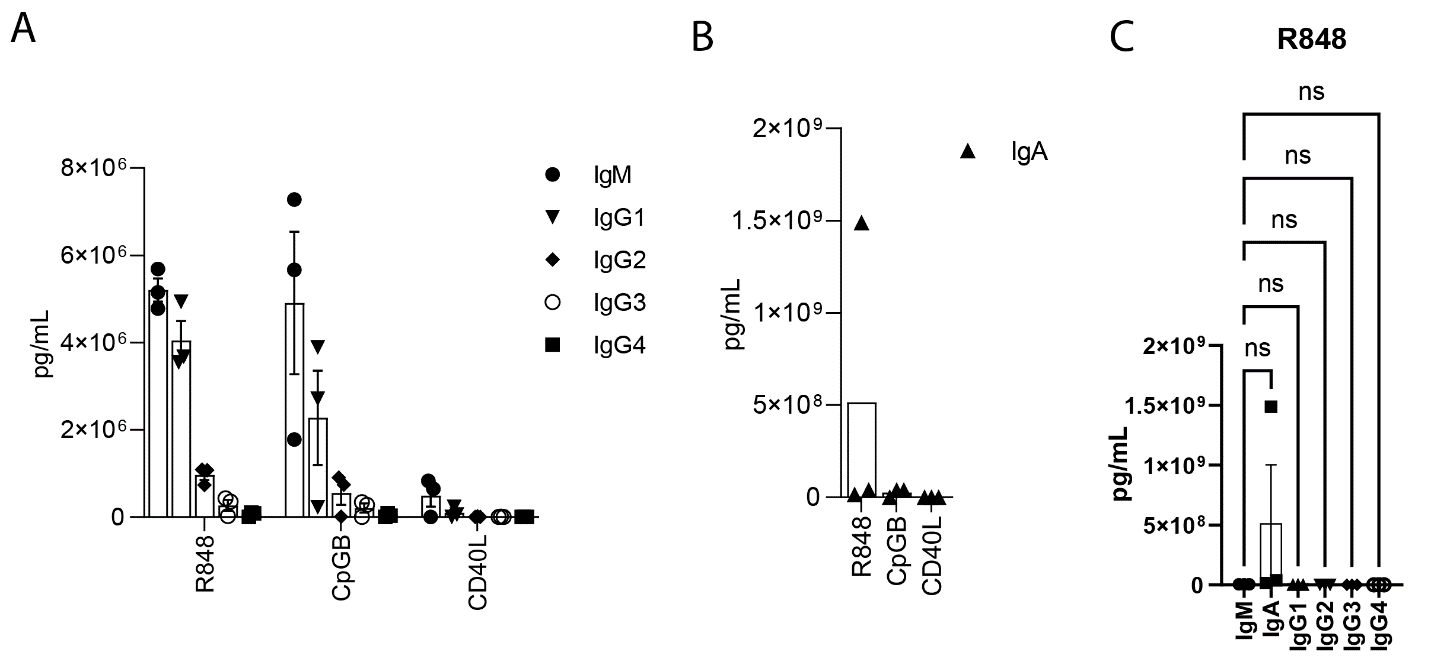

Fig. S16. Antibody response to positive control conditions from non-selective B cell stimulation. (A) Titres from Legendplex assay for positive controls R848, CpG, and recombinant CD40. Data points are B cells from independent donors (n=3 each condition) for IgM and IgG isotypes (IgA shown separately as very high values for one individual). (B) IgA (C) Combined data from A&B for R848.

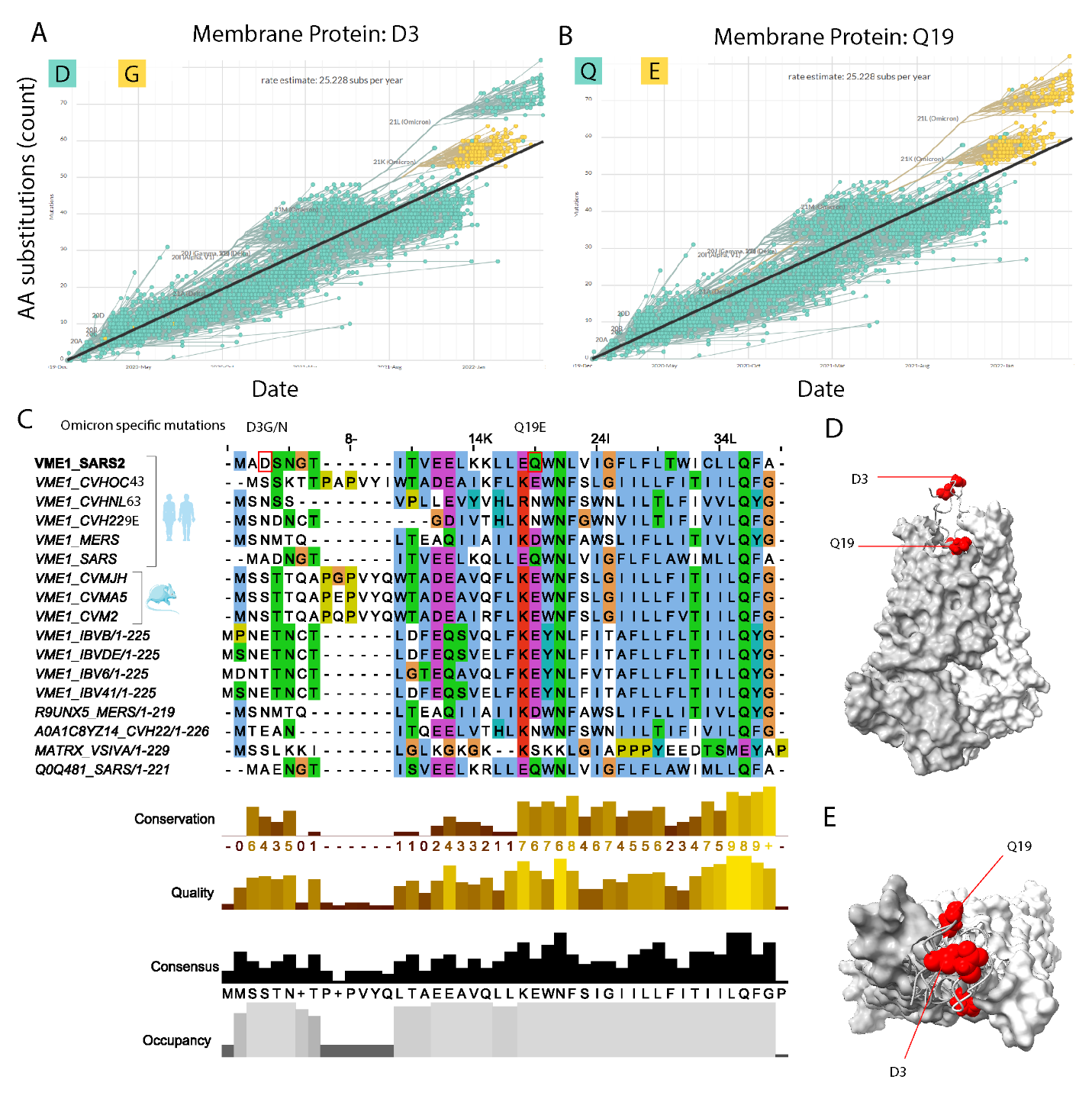
Fig. S197. Alignment of SARS-CoV-2 membrane protein with other coronaviruses. (A)

Phylogenetic diagram plotting number of non-synonymous mutations (y-axis) against date of sample for a sample of all public SARS-CoV-2 genomes. Each point is a SARS-CoV-2 sequence and colour represents the amino acid at position 3 or (B) amino acid at position 19. (data courtesy of <https://nextstrain.org/>, GISAID data.) (C) Multiple sequence alignment of various human and animal-infecting coronaviruses. (D) Position of the D3 and Q19 amino acids on the membrane protein dimer. The D3 position (which may be alternatively glycosylated in BA.5) is the most exposed position on the membrane protein dimer.

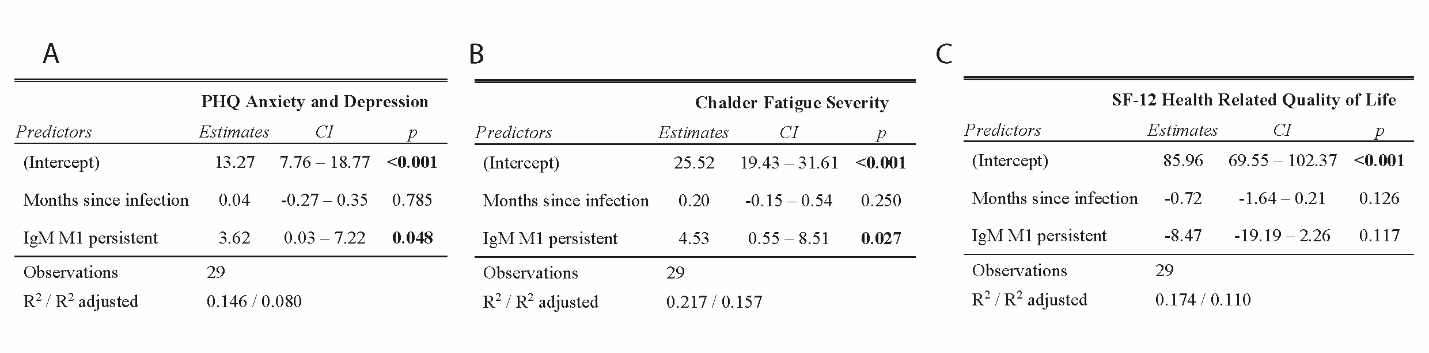

Fig. S18. Linear regression for long COVID cohort showing the predictive value of persistent IgM on anxiety and depression and Chalder Fatigue Severity. (A) PHQ anxiety and depression. (B) Chalder fatigue severity. (C) SF-12 health related quality of life.

Table S1. Manchester cohort subject characteristics

Data are median (IQR)m , where ^m^ is the number of missing data points, n (%) or n/N (%), where N is the total number with available data. PE, pulmonary embolism; IHD, Ischaemic Heart Disease; COPD, Chronic Obstructive Pulmonary Disease; HTN, Hypertension; PCR, Polymerase Chain Reaction; CRP, C-Reactive Peptide; BMI, Body Mass Index. ^a^Admission observations. *Subject died following a cerebrovascular event during admission. Missing data are missing as assay values reported as unavailable for these samples by the clinical team.

|  | **Total (36)** | **Ward based care (24)** | **Higher level care (12)** |
| --- | --- | --- | --- |
| **Age^a^** | 65 (53-73)^3^ | 65 (54.3-72.8) | 67 (51-76.5)^1^ |
| **Sex** |  |  |  |
| Female | 12/33 (36.3)^3^ | 8/22 (36.4)^2^ | 4/11 (36.4)^1^ |
| Male | 21/33 (63.6)^3^ | 14/22 (63.6)^2^ | 7/11 (63.6)^1^ |
| **BMI^a^** | 27.5 (25.2-30.5)^5^ | 27.9 (25.4-30.5)^4^ | 26.5 (25.0-30.8)^1^ |
| **Day of symptoms on recruitment** | 9 (6.5-11.5)^5^ | 10 (6.75-12)^4^ | 8 (5-10)^1^ |
| **Medical history** |  |  |  |
| Diabetes | 6/30 (20)^6^ | 3/20 (15)^4^ | 3/10 (30)^2^ |
| Asthma | 5/30 (16.7)^6^ | 5/20 (25)^4^ | 0/10 (0)^2^ |
| COPD | 5/30 (16.7)^6^ | 4/20 (20)^4^ | 1/10 (10)^2^ |
| Previous PE | 1/30 (3.3)^6^ | 1/20 (5)^4^ | 0/10 (0)^2^ |
| IHD | 3/30 (10)^6^ | 2/20 (10)^4^ | 1/10 (10)^2^ |
| HTN | 7/30 (23.3)^6^ | 5/20 (25)^4^ | 2/10 (20)^2^ |
| **Smoking status** |  |  |  |
| Never | 17/28 (60.7)^8^ | 12/18 (66.7)^6^ | 5/10 (50)^2^ |
| Ex-smoker | 10/28 (35.7)^8^ | 6/18 (33.3)^6^ | 4/10 (40)^2^ |
| Current | 1/28 (3.6)^8^ | 0/18 (0)^6^ | 1/10 (10)^2^ |
| **Chest imaging^a^** |  |  |  |
| Bilateral opacification | 27/33 (81.8)^3^ | 16/22 (72.7)^2^ | 11/11 (100)^1^ |
| Unilateral opacification | 4/33 (12.1)^3^ | 4/22 (18.2)^2^ | 0/11 (0)^1^ |
| Clear | 2/33 (6.1)^3^ | 2/22 (9.1)^2^ | 0/11 (0)^1^ |
| **Outcome** |  |  |  |
| Length of stay (days) | 7.5 (5.75 – 11.5) | 7 (5-10)^3^ | 9 (6.5-12)^1^ |
| Mortality | 7/32 (21.9) | 1*/21 (4.8)^3^ | 6/11 (54.5)^1^ |

Table S2. Manchester patient categorization information.

Type caption.

| **Severity** | **Criteria** |
| --- | --- |
| **Ward level care** | <10L or 60% FiO2 supplemental oxygen required to maintain oxygen saturations. |
|  | Managed in a ward-based environment. |
|  | Chronic NIV or CPAP (home use) or acute NIV for COPD. |
| **Higher level care** | Any of: |
|  | >10L or 60% supplemental oxygen required to maintain oxygen saturations. |
|  | Use of acute NIV (not for COPD). |
|  | Managed in ICU/invasive ventilation. |

Table S3. Oxford cohort subject clinical information.

Data are median (IQR)m , where^m^ is the number of missing data points, n (%) or n/N (%), where N is the total number with available data. PE, pulmonary embolism; IHD, Ischaemic Heart Disease; COPD, Chronic Obstructive Pulmonary Disease; HTN, Hypertension; PCR, Polymerase Chain Reaction; CRP, C-Reactive Peptide; BMI, Body Mass Index. ^a^Admission/Recruitment observations. Missing data are missing as assay values reported as unavailable for these samples by the clinical team.

|  | **Total** | **Asymptomatic/Mild (45)** | **Severe/Critical (25)** |
| --- | --- | --- | --- |
| **Age^a^** | 45.5 (32.8-57.0)^0^ | 38.9 (30.0-54.0)^0^ | 65.0 (48.0-78.0)^0^ |
| **Sex** |  |  |  |
| Female | 45/72 (62.5)^0^ | 35/45 (77.8)^0^ | 9/25 (36.0)^0^ |
| Male | 27/72 (37.5)^0^ | 10/45 (22.2)^0^ | 16/25 (64.0)^0^ |
| **Day pso/PCR+** | 40 (29-54.5)^0^ | 31.0 (28.0-56.0)^0^ | 43.0 (40.0-53.0)^0^ |
| **Jenner anti-spike (AU)** |  |  |  |
| IgG | 565.2 (251.3-2038.3)^14^ | 343.5 (184.4-813.8)^1^ | 3820 (2268-5921)^13^ |
| IgA | 81.1 (45.8-320.5)^33^ | 64.7 (41.9-109.2)^12^ | 392.4 (292.8-447.5)^21^ |
| IgM | 117.4 (35.2-471.0)^33^ | 87.6 (28.9-271.4)^12^ | 758.9 (374.1-1070.9)^21^ |
| **Monogram pseudoneutralising antibodies (IC50)** | 838.1 (314.5-3090.8)^9^ | 388.7 (198.7-868.7)^3^ | 3367 (1944-10102)^4^ |

Table S4. Oxford cohort categorization information.

Severity was graded as a mild, severe or critical according to World Health Organization guidelines. One subject had disease severity graded as moderate as they did not meet other criteria for severe/critical infection but was included with this group on the basis of requiring hospitalisation and supplemental oxygen support. Severe and critical groups were combined in all analyses.

| **Severity** | **Criteria** |
| --- | --- |
| **Asymptomatic/Mild** | Non-hospitalised for COVID-19. |
|  | None of the criteria for severe/critical classification. |
| **Severe/Critical** | At least one of: |
|  | Respiratory rate > 30/min. |
|  | Blood oxygen saturation <90%. |
|  | Arterial oxygen partial pressure (PaO2)/fraction of inspired O2 (FiO2) < 300 mmHg. |
|  | Respiratory failure requiring mechanical ventilation. |
|  | Shock or other organ failure requiring ITU admission. |

Table S5. Edinburgh longitudinal cohort subject clinical information.

Data are median (IQR)m , where^m^ is the number of missing data points, n (%) or n/N (%), where N is the total number with available data. Abbott anti-N (anti-Nucleoprotein IgG), Diasorin S1/2 (anti-spike S1/2 IgG) ; Siemens RBD (COV2T RBD total); RocheN (Nucleoprotein total), RocheS (RBD total); Diasorin trispike (S stabilized trimeric spike protein IgG); Euroimmun (spike S1’ subunit IgG); Abbott IgGII Quant (RBD IgG); PCR, polymerase chain reaction; Pseudovirus neutralization assay uses SARS-CoV-2 Wuhan-Hu-1 pseudovirus. ^a^Admission/Recruitment observations. Missing data are missing as assay values reported as unavailable for these samples by the clinical team.

|  | **Visit 1 (111)** | **Visit 2 (92)** | **Visit 3 (54)** | **Visit 4 (4)** |
| --- | --- | --- | --- | --- |
| **Age** | 47.0 (32.5-55.0)^0^ | 48.5 (33.8-56.3)^0^ | 49.0 (29.5-55.8)^0^ | 48 (42.3-51.0)^0^ |
| **Sex** |  |  |  |  |
| Female | 80/111 (72.1)^0^ | 63/92 (68.4)^0^ | 36/54 (66.7)^0^ | 2/4 (50)^0^ |
| Male | 31/111 (27.9)^0^ | 29/92 (31.5)^0^ | 18/54 (33.3)^0^ | 2/4 (50)^0^ |
| **Day PCR+** | 42 (34-46)^0^ | 42 (34-46)^0^ | 66 (61.3-70.0)^0^ | 95.5 (93-97)^0^ |
| **Hospitalised** | 5/111 (4.5)^0^ | 4/92 (4.3)^0^ | 2/54 (3.7)^0^ | 0/4 (0)^0^ |
| **Commercial assay (AU)** |  |  |  |  |
| Abbott anti-N | 4.74 (2.82-6.43)^0^ | 4.48 (2.43-6.52)^0^ | 4.67 (2.27-6.42)^0^ | 4.92 (1.70-8.26)^0^ |
| Diasorin S1/2 | 62.5 (29.3-103.0)^0^ | 64.9 (34.7-111)^0^ | 64.9 (26.9-119)^0^ | 60.8 (25.2-94.6)^0^ |
| Siemens RBD | 16.0 (5.5-46.1) | 19.5 (8.81-49.0)^0^ | 24.1 (9.4-62.1)^0^ | 32.7 (6.2-61.4)^0^ |
| RocheN | 40.8 (14.8-82.1) | 55.2 (23.4-97.6)^0^ | 63.7 (20.6-105)^0^ | 57.0 (27.2-84.1)^0^ |
| RocheS | 93.4 (21.7-236.0)^0^ | 105 (43.9-245)^0^ | 142.5 (44.8-266)^0^ | 169 (43.1-322)^0^ |
| Diasorin trispike | 113 (46.7-217)^0^ | 98.5 (44.0-188)^0^ | 94.3 (35.2-180)^0^ | 59.4 (34.9-106)^0^ |
| Euroimmun | 3.9 (2.2-6.0)^0^ | 4.0 (2.3-6.0)^0^ | 3.8 (1.7-6.0)^0^ | 3.3 (1.7-4.8)^0^ |
| SiemensCOVG | 8.07 (3.08-21.0)^0^ | 7.54 (3.05-17.7)^0^ | 7.0 (3.0-17.2)^0^ | 5.32 (2.26-9.33)^0^ |
| Abbott IgG Quant | 846 (333-2024)^0^ | 665 (290-1724)^0^ | 665 (254-1634)^0^ | 653 (291-1117)^0^ |
| **Pseudovirus**  **neutralising antibodies (IC50)** | 323 (152-679)^11^ | 259 (124-510)^0^ | 198 (74.3-361)^0^ | 116 (81.7-193.5)^0^ |

Table S6. SNBTS cohort subject characteristics

Data are median (IQR)m , where^m^ is the number of missing data points, n (%) or n/N (%), where N is the total number with available data. ^a^Recruitment observations.

|  | **SNBTS (200)** |
| --- | --- |
| **Age^a^** | 48 (35-55.3)^0^ |
| **Sex** |  |
| Female | 99/200 (49.5)^0^ |
| Male | 101/200 (50.5)^0^ |
| **EuroImmun spike IgG titre (AU)** | 4.45 (2.00-6.60)^0^ |

Table S7. Participant characteristics for in vitro B cell stimulation experiments

| **Participant** | **Condition** | **Participant** | **Ethnicity** | **Sex** | **Age** | **COVID vaccine history** |
| --- | --- | --- | --- | --- | --- | --- |
| N-01 | COVID-naive | N-01 | Indonesian | M | 40 | Sinovac x2 |
| N-02 | COVID-naïve | N-02 | Chinese | M | 24 | Comirnaty x3 |
| N-03 | COVID-naive | N-03 | Indian | F | 32 | Pfizer x4 |
| P-01 | 8 weeks post-molecular test confirmed SARS-CoV-2 breakthrough infection | P-01 | White British | F | 30 | Pfizer x3 |
| P-02 | 6 weeks post-molecular test confirmed SARS-CoV-2 breakthrough infection | P-02 | White British | M | 45 | Pfizer x3 |
| P-03 | 10 weeks post possible COVID breakthrough infection. Confirmed exposure, mild symptomatic illness, no confirmatory test. | P-03 | White British | M | 50 | Pfizer x3 |

Table S8. Long COVID cohort subject characteristics

Data are median (IQR)m , where^m^ is the number of missing data points, n (%) or n/N (%), where N is the total number with available data. One missing data point as clinical information not received for this sample (excluded from analyses as possible sample labelling error). ^a^Recruitment observations.

|  | Long COVID cohort (30) |
| --- | --- |
| **Age^a^** | 42 (35-49)^1^ |
| **Sex** |  |
| Female | 19/29 (65.5)^1^ |
| Male | 10/29 (34.5)^1^ |
| **Months since infection** | 16 (14-18)^1^ |
| **PHQ** | 15 (12-18)^1^ |
| **Chalder Fatigue Severity** | 30 (27-34)^1^ |
| **FCS12** |  |
| Combined | 72.5 (63.2-76.3)^1^ |
| PCS | 33.1 (26.2-41.6)^1^ |
| MCS | 35.2 (24.7-50.0)^1^ |

Table S9. Eight prioritized immunodominant epitopes

| Protein | Peptide | Start | End | Peptide sequence |
| --- | --- | --- | --- | --- |
| Membrane | M1 | 1 | 19 | MADSNGTITVEELKKLLEQ |
| Nucleoprotein | N2 | 14 | 48 | RITFGGPSDSTGSNQNGERSGARSKQRRPQGLPNN |
|  | N11 | 160 | 171 | QLPQGTTLPKGF |
|  | N14 | 192 | 227 | NSSRNSTPGSSRGTSPARMAGNGGDAALALLLLDRL |
|  | N28 | 380 | 419 | QALPQRQKKQQTVTLLPAADLDDFSKQLQQSMSSADSTQA |
| Spike | S39 | 552 | 587 | LTESNKKFLPFQQFGRDIADTTDAVRDPQTLEILDI |
|  | S51 | 806 | 825 | LPDPSKPSKRSFIEDLLFNK |
|  | S67 | 1139 | 1169 | DPLQPELDSFKEELDKYFKNHTSPDVDLGD |

Data S1. (separate file)

Zipped tab-separated values file detailing all 196 peptides investigated in this study.

Data S2. (separate file)

Zip of tab-separated values files with comprehensive dataset of ELISA results for individual samples used in analyses in this paper.

Data S3. (separate file)

Zip of tab-separated values files with calculated SASA for all possible spike, envelope, membrane, and nucleoprotein peptides 10-100aa.

**Data S4 (separate file)**

Large image file. High resolution low-microscopy photographs of B cells from individuals recently recovered from SARS-COV-2 breakthrough infection, stimulated with VLPs and control conditions in the presence of cytokines at day 8 and day 14 post culture.
